# Deep learning for interactive and automated inner retinal layer segmentation in OCT of patients with retinitis pigmentosa using limited training data

**DOI:** 10.64898/2026.06.16.26355668

**Authors:** Dorothea Laurence, Martin Schilling, Nina-Antonia Grimm, Emilie Macé, Sebastian Bemme, Constantin Pape

**Author notes:** Constantin Pape and Sebastian Bemme contributed equally as senior authors and share last authorship. Funding disclosures: none. Commercial relationship disclosures: none.

## Abstract

**Purpose:** New therapeutic strategies such as optogenetics have created a need for accurate tracking of inner retina degeneration in Retinitis pigmentosa (RP) patients. We introduce two tailored deep learning models to segment the RNFL (retinal nerve fibre layer), GCIPL (ganglion cell inner plexiform layer), INL (inner nuclear layer), CFT (central foveal thickness) and RPE (retinal pigment epithelium) in RP: The first is based on a Segment Anything Model (SAM), the second on nnU-Net. To our knowledge, SAM has not yet been applied to retinal layers in OCT data.

**Methods:** SD-OCT images of a retrospective cohort of 37 RP patients were included. Data for four training cycles were prepared semi-automatically in MATLAB, then assessed and corrected by three expert graders. 1,700 segmented B-Scans from two open datasets were used for pretraining. For post-processing, semantic retinal boundary detection was developed. The final models, OCT-SAM and nnU-Net, were trained on 228 annotated RP scans. Detected layer thicknesses were validated against manual segmentation at 90 random points in 30 OCT B-Scans. Finally, OCT-SAM was tested on three RP cases with retrospective, longitudinal OCT data.

**Results:** nnU-Net achieved a precision, recall and F-1 score of 0.96 while OCT-SAM performance resulted in slightly lower values of 0.93, 0.8 and 0.85, respectively. OCT-SAM measurements had low bias and good agreement with manual annotations, confirming reliability.

**Conclusions:** OCT-SAM enabled fast data annotation and tool integration, whereas nnU-Net provided the best segmentation performance. OCT-SAM demonstrated longitudinal reproducibility and detected RP-characteristic pathologies and degenerative changes. Future work will extend OCT-SAM to 3D OCT segmentation.

## Introduction

Retinitis pigmentosa (RP), a group of inherited retinal diseases (IRDs), is the leading cause of visual impairment and blindness in the non-elderly (below the age of 60 years), with an average age of diagnosis of 35.1 years ^1^. The prevalence of RP varies between countries and is estimated at 1 in 4000 globally. Up to date, more than 3000 mutations have been identified as its potential cause ^2^. RP presents itself with a loss of rod photoreceptors followed by cone photoreceptor degeneration. Outer retinal layers are affected in sequence with inner retinal layers, including the retinal nerve fiber layer (RNFL), retinal ganglion cells (RGCs) and bipolar cells (BPCs).

While current treatment options are limited to carriers of one specific mutation ^3^, novel therapeutic strategies such as optogenetics, targeting inner retina cells, have made promising advances^4^. These developments have given rise to a new need for accurate tracking of the degenerative processes of the inner retina in RP patients. Spectral domain and swept source optical coherence tomography (SS- or SD-OCT) scans provide insight into RGCs’ soma and axon degeneration by measuring the thickness of the ganglion cell inner plexiform layer (GCIPL) ^5–7^ and the RNFL ^5,8^. BPCs’ soma changes can be detected in the inner nuclear layer (INL)^6^. Automated OCT segmentation tools by OCT-manufacturers such as Heidelberg Engineering, however, fail to correctly segment degenerate retinal layers in diseases such as RP due to a loss of key boundaries found in healthy retinae ^6,9^. Therefore, more robust alternatives such as deep learning algorithms have begun to play an increasingly important role for disease identification and monitoring via those retinal imaging techniques ^10^.

Current applications of deep learning are automated segmentation of individual retinal layers, identification and quantification of disease-specific biomarkers for disease identification, monitoring, and prognosis ^11,12^. While there are well-developed algorithms for common eye diseases such as diabetic retinopathy, age related macular degeneration, and glaucoma, deep learning models for IRDs, specifically RP, are rare. One reason for this is the scarcity of available IRD datasets ^13^. Other challenges include the common motion artifacts and reduced image quality in IRDs caused by fixation deficits of patients ^14^.

Deep learning OCT segmentation developed to date specifically for IRDs and RP focused on the assessment of photoreceptor integrity and related retinal layers such as the ellipsoid zone (EZ), the presence of the retinal pigment epithelium (RPE) and the outer nuclear layer (ONL). ^13,15–18^ To our knowledge, however, deep learning has not yet been applied to identify and monitor inner retina layers such as the RNFL, the GCIPL and the INL in optical coherence tomography images of RP patients.

In preparation for our “Prospective exploratory cohort study on ganglion cell degeneration in Retinitis pigmentosa patients” (ClinicalTrials.gov ID: NCT07056738), we thus introduce two tailored deep learning models to segment the inner retina layers of RP patients and track their changes over time. Our models are based on two state-of-the-art architectures. The first, which we call OCT-SAM, is based on MedicoSAM ^19^, an extension of the Segment Anything Model ^20^ (SAM) for medical imaging. SAM is a vision foundation model for image segmentation tasks that enables interactive and automatic segmentation in a wide range of imaging modalities. It has been applied and improved for medical imaging tasks in many studies ^21,22^. However, to the best of our knowledge, SAM has not yet been applied to analyse retinae in OCT data. Second, we also train a model using the nnU-Net v2 architecture, which is the most established model for automatic semantic segmentation in medical imaging. These models have complementary strengths: OCT-SAM enables fast data annotation and tool integration, whereas nnU-Net provides the best automatic segmentation quality.

## Methods

### Study population and image acquisition

This study was approved by the University Medical Center Göttingen Institutional Ethics Committee (application no., 13/1/25) and adheres to the tenets of the Declaration of Helsinki. For the study, we included a retrospective cohort of 31 RP patients presenting at the Department of Ophthalmology, University Medical Center of Göttingen, between 2019 and 2025. Inclusion criteria were a genetic and/or clinical diagnosis of RP, image acquisition via Spectralis SD OCT (Heidelberg Engineering) and sufficient image quality to differentiate individual inner and outer retinal layers. SD-OCT-scans were chosen to cover a variety of disease stages and complications such as the presence of gliosis, cystoid macular edema (CME) and central RPE atrophy (tables 1-4).

### Segmentation and annotations for training cycles

Initially, layer boundaries of 96 selected B-Scans (Tables 1 and 5) were first segmented semi-automatically using a Canny edge detector (modified to only consider vertical intensity gradients), then manually corrected using MATLAB (MathWorks) by an experienced first grader and validated by an expert second grader. In cases lacking consensus, an expert third grader was consulted for final validation.

We segmented the following layers: (1) RNFL; (2) GCIPL; (3) INL; (4) outer plexiform layer (OPL); (5) layer between the inner border of the outer nuclear layer (ONL) and the outer border of the myoid zone (MZ); (6) EZ; (7) layer between the outer border of the EZ and the outer border of the RPE-Bruch-Membrane-complex (RPEBM) (see Figure 1, left image). Additionally, we classified the stage of RP using the staging method of Oh et al.^23^: an early stage was defined by a width of the preserved EZ-zone of ≥ 2500 μm, a moderate stage was defined as < 2500 μm EZ width, and an advanced stage by the absence of the EZ.

**Figure 1:**
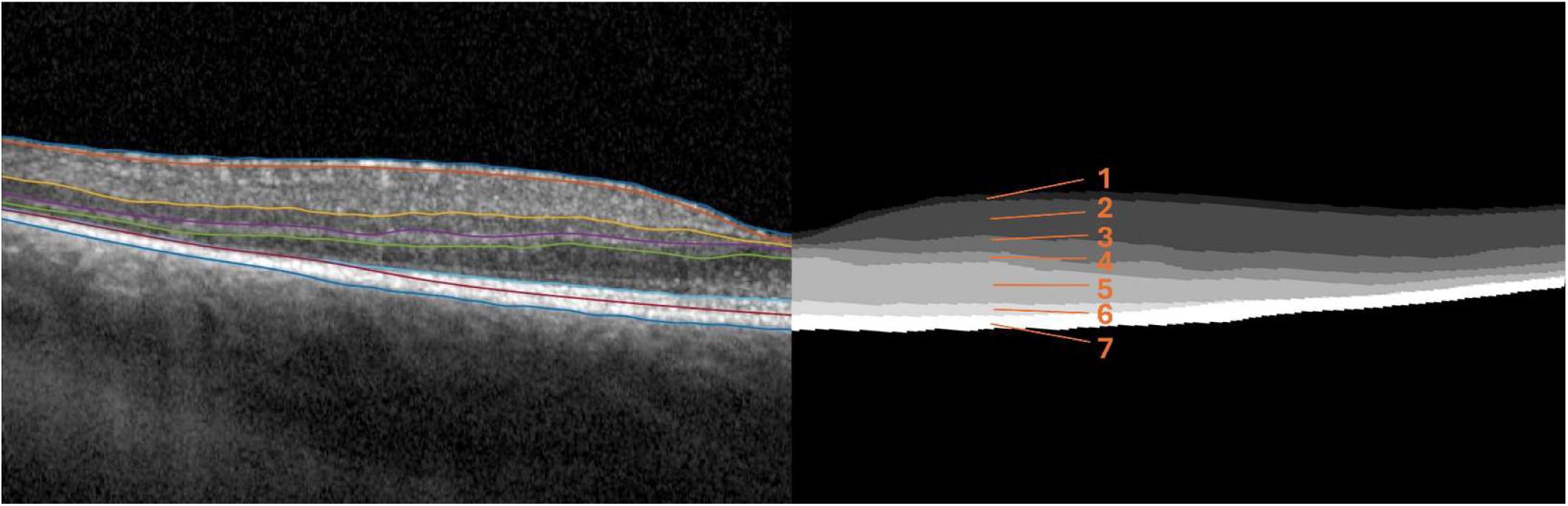
Segmented B-Scan (left) with converted semantic layer masks (right). 1-7 = segmented layers. 1 = RNFL; 2 = GCIPL; 3 = INL; 4 = OPL; 5 ONL and outer border of MZ; 6 = EZ; 7 = RPEBM.

The segmented B-Scan images were then converted into semantic layer masks via MATLAB serving as ground truth for the training of the model (see Figure 1, right image). We used 76 of the B-Scans to train the initial version of the model (next section) and held out 20 as a separate test set.

For the second, third and fourth training cycle, selected B-Scans (Tables 2-4 and 6-8) were pre-segmented with OCT-SAM and manually corrected using the interactive napari plugin (next section). The model improved with every training iteration, thus leading to faster data annotation.

### Segmentation methodology

We adapted the SAM ^20^ for interactive and automatic segmentation of retinal layers in B-scans. To this end, we used the architectural changes of μSAM ^22^, which introduced a convolutional segmentation decoder that predicts foreground probabilities and normalized distances to object boundaries and centroids for the purpose of automatic instance segmentation. We initialized the model with the pretrained weights of MedicoSAM ^19^, an extension of μSAM for medical imaging. We then further this model on an initial dataset of 1,700 B-Scans with annotations for individual retinal layers from two prior studies ^24,25^, using the training implementation of μSAM. Notably, this training procedure improves the model’s capability for interactive segmentation (via the SAM architecture) and for automatic segmentation (via μSAM’s segmentation decoder). We then trained this model on the 76 scans of the initially annotated B-Scans of RP patients; with 20 scans held out as independent test data. This model was then used to interactively annotate the training data for the next cycles (see previous section), followed by re-training after each cycle. The final model — called OCT-SAM — was trained on a total of 228 annotated scans.

After training, OCT-SAM accurately segmented individual retinal layers based on point or box prompts. Its segmentation decoder predicted the foreground area and the normalized distance to the nearest layer boundary. Based on these outputs, we developed a method for automatic instance segmentation, i.e. segmenting all retinal layers in a B-Scan without user input. To this end, we identified likely boundaries as areas in the prediction with a normalized distance smaller than 0.5, then computed a directed vertical distance transform to this area, normalized these distances within a sliding window of 255 x 1 pixels, found connected areas with a value larger than 0.6, and used the centroids of these areas as point inputs for OCT-SAM’s prompt encoder. We applied non-maximum suppression to the predicted masks to retain only likely retinal segments. We found this procedure superior to watershed-based segmentation approaches that did not employ the promptable segmentation feature. It is similar to the automatic prompt generation method for automatic cell segmentation described in (Archit and Pape 2026). We then implemented a further post-processing logic that takes the organization of retinal layers into account. Firstly, the prediction is scanned to identify retinal layers that span the entire field of view horizontally. Secondly, the other retinal layers are merged based on their position in relation to these layers. Finally, instances thinner than 5 pixels are removed to avoid prediction artifacts.

In addition, we trained a nnU-Net v2, first training it on the public data and then fine-tuning it on our annotated data. This model directly addresses semantic segmentation, i.e. it directly predicts the layer identities. Unlike OCT-SAM, it does not support interactive data annotation and also does not provide direct tool integration (see next section).

### Automatic measurements & segmentation tool

We implemented functionality for measuring the following parameters of retinal layers based on their segmentation:

- Area
- Horizontal extension
- Minimal thickness
- Maximal thickness
- Mean thickness
- Thickness standard deviation

We then integrated the segmentation and measurement functionality into the napari-based GUI of μSAM, enabling the segmentation and analysis of individual B-Scans extracted from an OCT. Note that this tool offers automatic segmentation, interactive segmentation, and manual correction of segmentation layers, enabling efficient and convenient correction of automatic segmentation results (if needed). An overview of this tool is shown in Figure 2.

**Figure 2:**
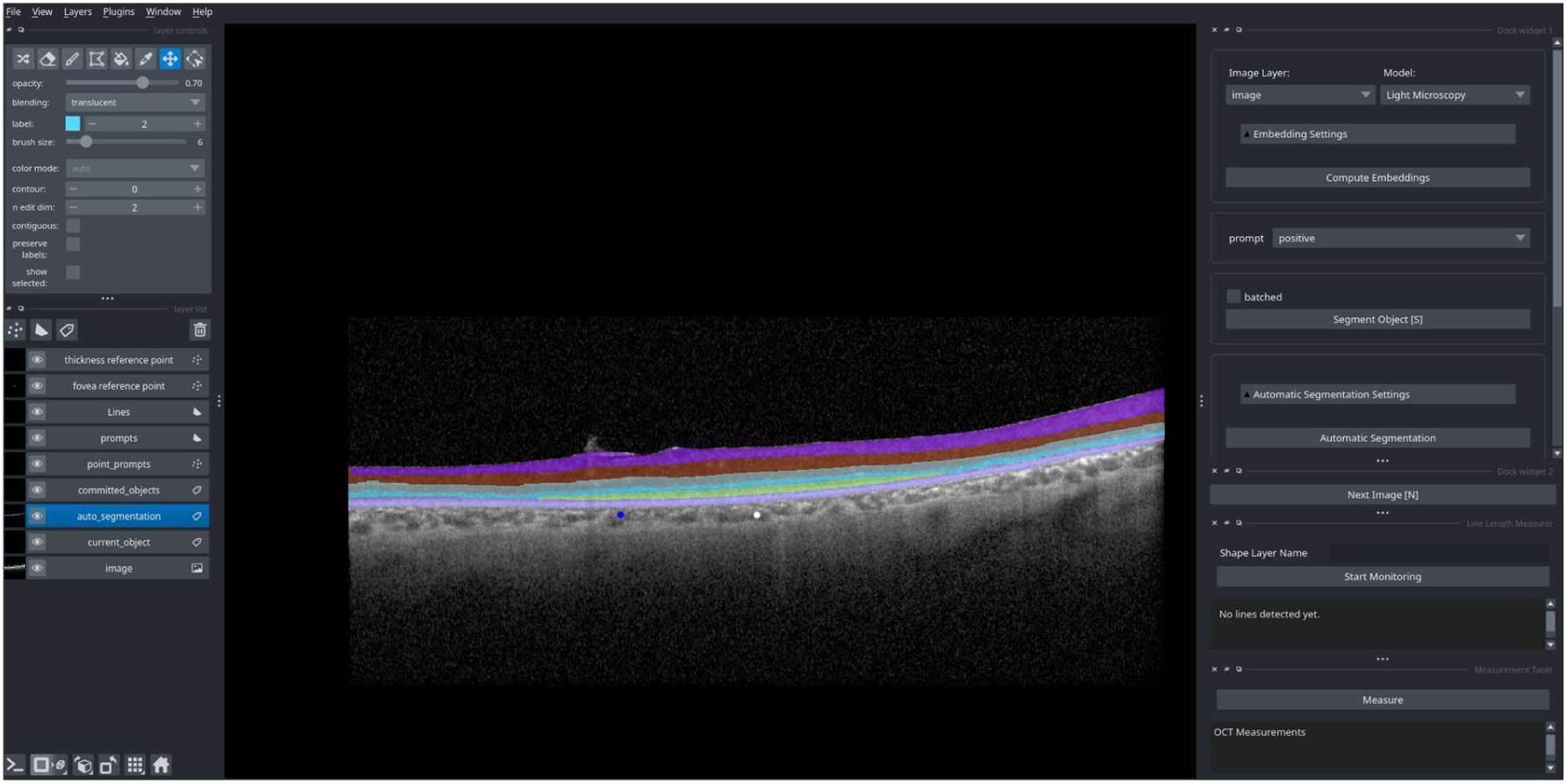
The user interface of our napari plugin for interactive and automatic segmentation of retinal layers as well as derived layer measurements.

Our software is well documented and available at https://github.com/computational-cell-analytics/oct-sam. Instructions for downloading the OCT-SAM model and the nnU-Net model are available in the repository.

## Results

### Accurate segmentation of retinal layers in RP patients

We evaluated the two models for retinal layer segmentation, OCT-SAM and the nnU-Net, on held out test data comprising 20 B-Scans with annotations (Table 1 and 5). Examples for manual segmentations, conversion into ground truth masks and outputs of both OCT-SAM and nnU-Net are shown in Figure 3. Figure 4 shows different metrics (Dice Score, Precision, Recall, F1-Score; higher is better for all) that evaluate the respective results compared to the gold standard annotations. Here, we evaluate two segmentation strategies for OCT-SAM, using the default μSAM segmentation strategy (OCT-SAM*) and the custom strategy we developed for the retinal layer segmentation (OCT-SAM, see Methods). The custom strategy is substantially better. The nnUNet is the most accurate method overall, slightly better than OCT-SAM. The nnUNet is the most accurate method overall, slightly better than OCT-SAM.

**Figure 3:**
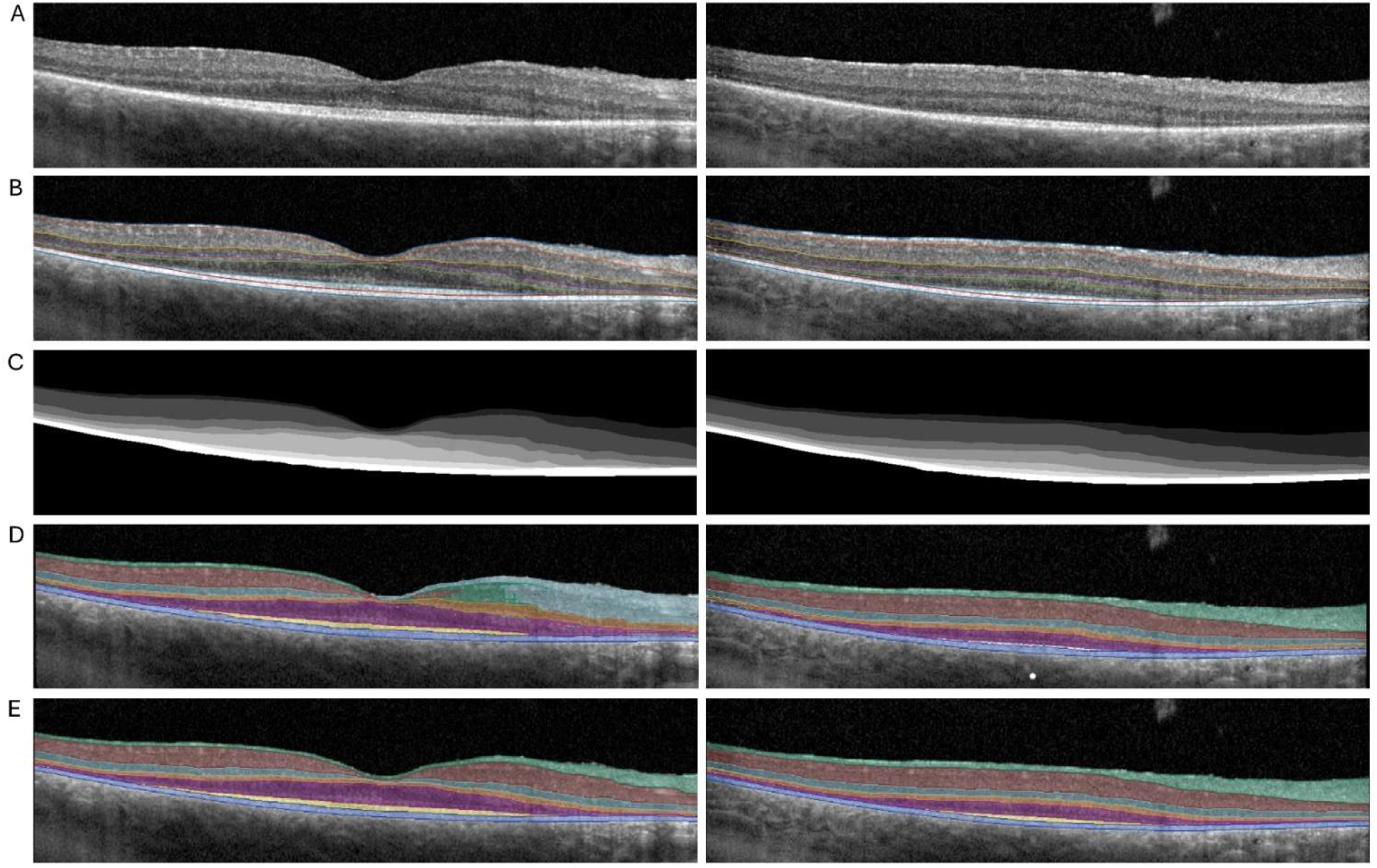
Segmentation results OCT-SAM (left column) vs. nnU-Net (right column) on two B-Scan-images of the same volume scan. A = original B-Scan image. B = manual segmentation. C = ground truth mask. D = automatic OCT-SAM output. E = automatic nnU-Net output. Layers in D and E: RNFL = turquoise, GCIPL = brown, INL = light blue, OPL = orange, ONL = purple, EZ = yellow, RPE = dark blue.

**Figure 4:**
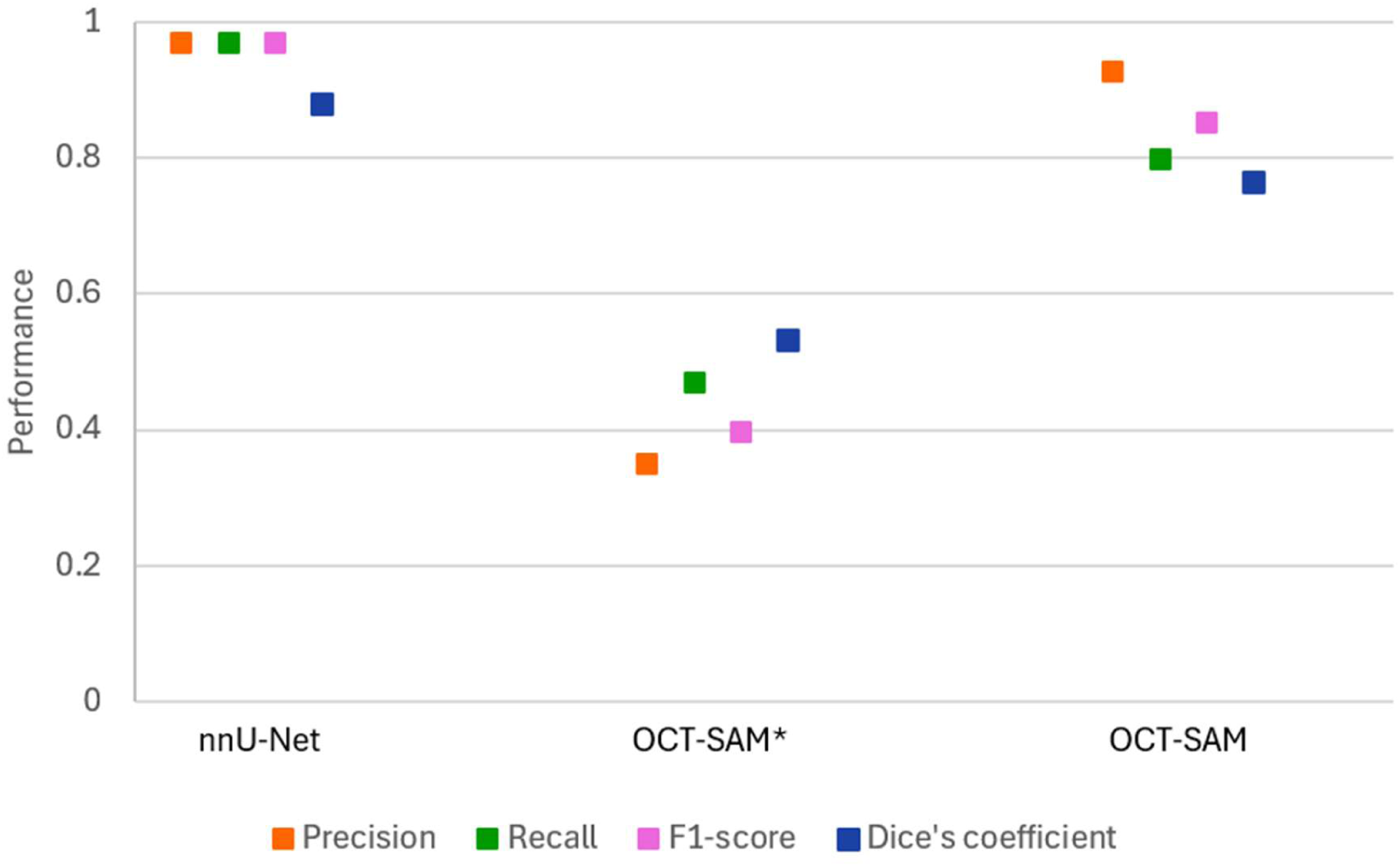
Evaluation of OCT-SAM and nnU-Net, trained on our complete training set (n = 228), on the test set (n = 20). The models were pretrained on publicly available OCT data (n = 1,700). For OCT-SAM we evaluated two segmentation methods, the default segmentation approach from μSAM (OCT-SAM*) and the custom approach described in the Methods section (OCT-SAM). Higher values are better for all metrics.

Furthermore, we investigated the impact of the amount of training data on model performance. Figure 5 compares OCT-SAM and nnU-Net that were trained exclusively on the publicly available data (public) with versions that were trained on subsets of our training data, using 1, 5, 10, 25, 50, 100, and all images. The respective models were initialized with the pretrained weights of the public model. We chose this set-up to study data annotation in multiple cycles systematically. The pre-trained version of OCT-SAM performs slightly better than the corresponding nnU-Net. Both models improve substantially when training on our task-specific data. The performance increases substantially with only a few images (5-10). Performance improvements are slightly lower and plateau earlier for OCT-SAM, leading to an advantage of nnU-Net when trained on many images.

**Figure 5:**
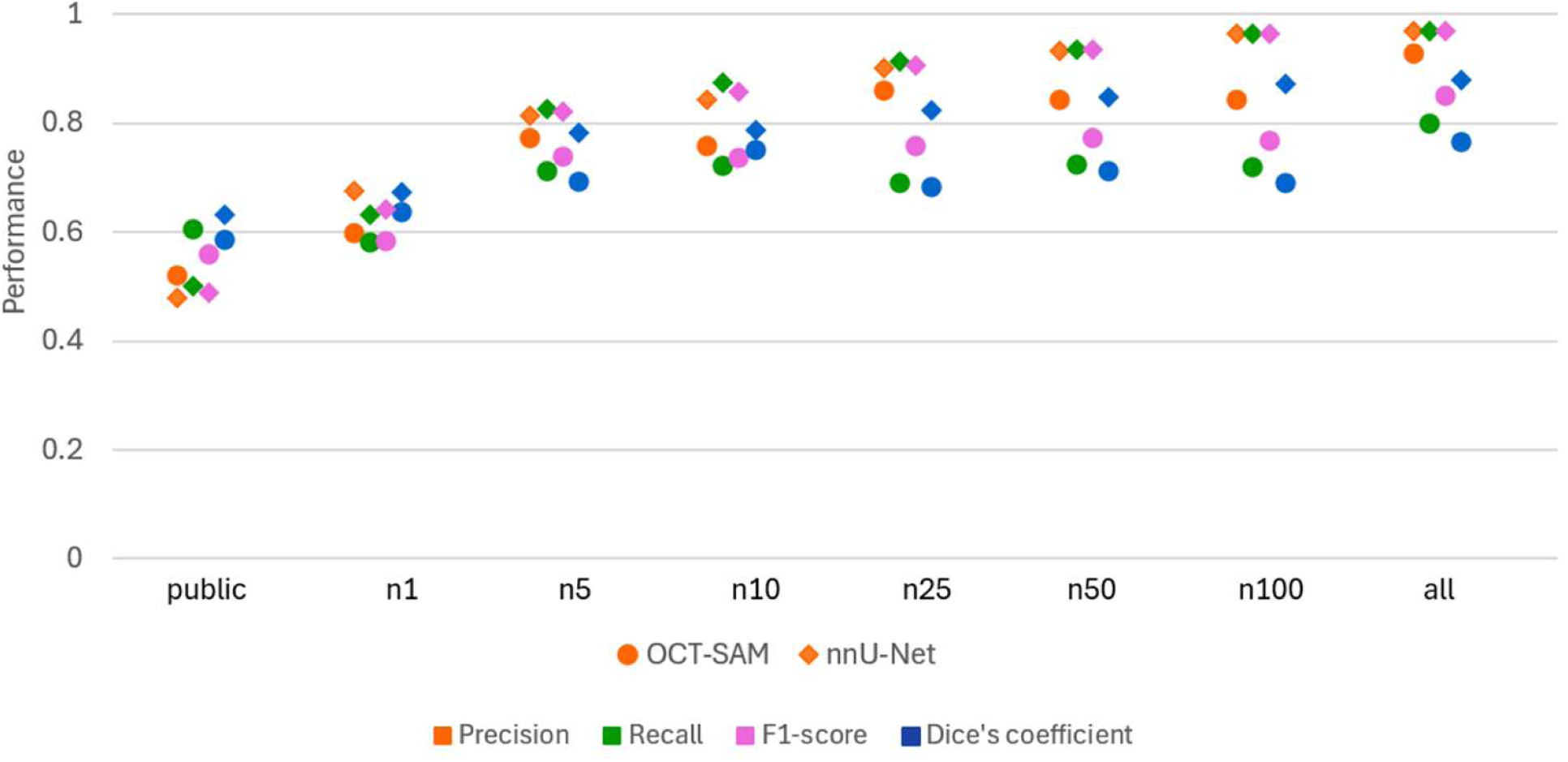
Evolution of segmentation performance with increasing number of training images. We evaluated OCT-SAM and nnU-Net that were trained on publicly available data (public) and on 1, 5, 10, 25, 50, 100, and all (n = 228) images from our training set. The evaluation was done on our held-out test set (n = 20) following the same approach as before.

The previous experiment clearly showed the need for training on specific data, motivating the integration of interactive data annotation within our tool in order to speed up the otherwise laborious data annotation process. Hence, we also studied OCT-SAM for interactive segmentation, using the same models as in the previous experiment and the evaluation strategy from Archit at al. ^22^ that derives prompts for interactive segmentation from the annotations and then evaluates the predicted masks. Figure 6 shows the corresponding evaluation for the different retinal layers and the corresponding average scores, for interactive segmentation with 1 - 4 point prompts. Note that we evaluated this model in a more challenging setting, where each layer was segmented individually. Thus, the scores are not directly comparable to other experiments where we evaluated the overall segmentation. Overall, we see that interactive segmentation with OCT-SAM improves substantially with additional training data, in particular for difficult to segment layers.

**Figure 6:**
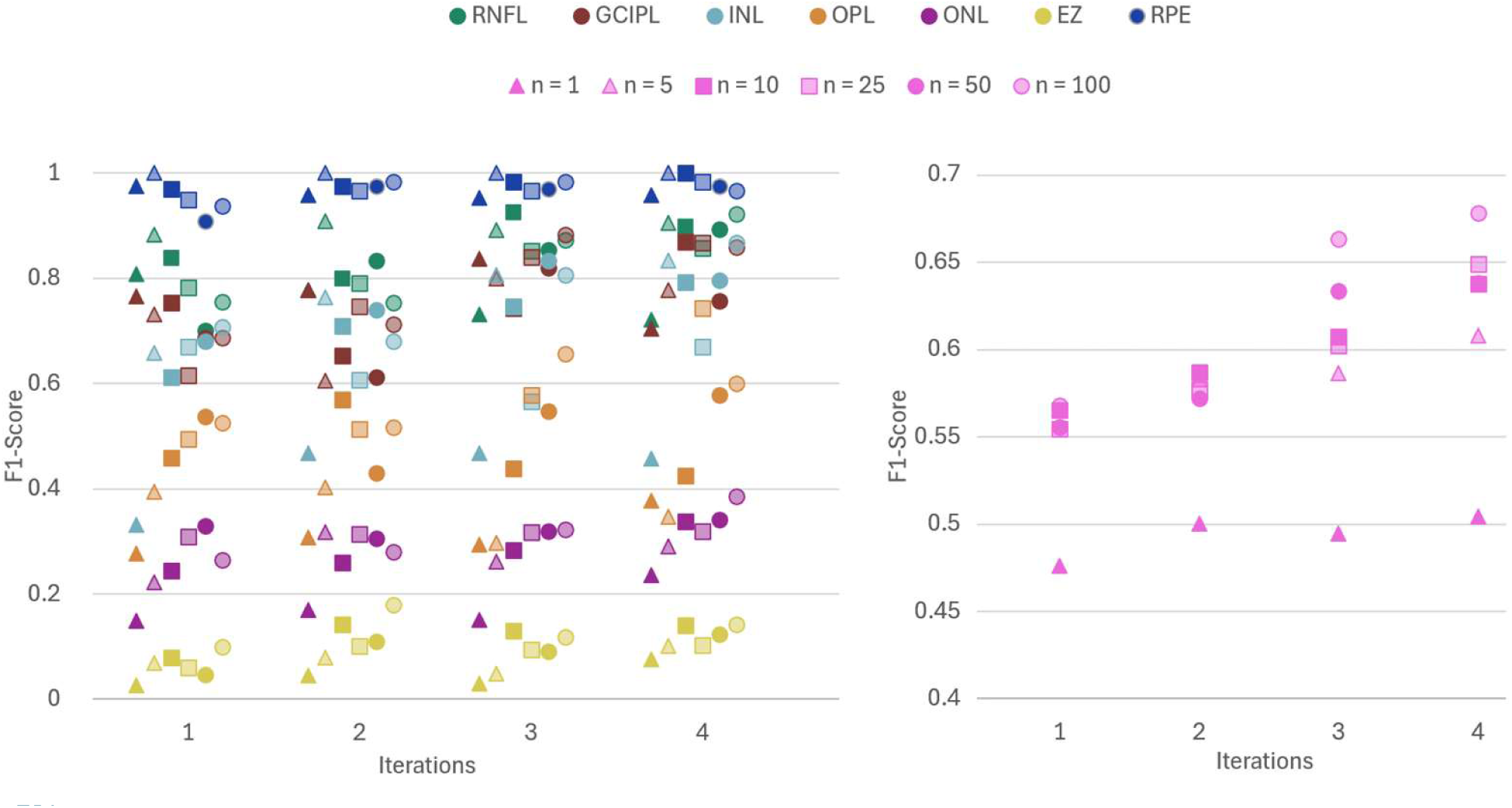
Evaluation of interactive segmentation performance with OCT-SAM with increasing number of training images. Here, the model is evaluated by automatically deriving 1-4 point locations from the respective annotation mask per retinal layer, prompting the model with this point(s) and evaluating its prediction against the annotation. For settings with multiple prompts (2-4), the points are derived iteratively from previous errors in the prediction in order to simulate. The left plot shows the F1-Scores for individual layers, right shows the average over layers.

### Accurate retinal layer segmentation enables the automatic measurement of key characteristics of retinal layers

To validate the accuracy of the automatic measurement feature of OCT-SAM, we established a new dataset of patients and corresponding scans not previously used for either training or validation (Table 9). Following along our method for the training and evaluation data, we utilized raw-OCT images, subsequently converted to .tif files. We randomly selected 5 OCT-B-Scans of each patient, and then for each randomly selected 3 different positions on the horizontal axis, resulting in 90 measuring points for validation (Table 10). The positions were marked on the B-Scans via color-coded points. We applied the automatic measurement tool at these positions, yielding the layer thicknesses of the RNFL, GCIPL, INL, OPL, ONL, EZ and RPE. Additionally, we used the “central (foveal) thickness” (CFT) automated measurement feature, assessing the combined retinal thickness at the center of each scan. An expert grader measured the corresponding thicknesses at each marked position manually within the Heidelberg program. We calculated the absolute difference, standard deviation and percentage difference between the measurements of the segmentation tool and the manual results to quantify the magnitude and relative impact of measurement deviations (Table 11). These differences were also assessed using Bland-Altmann plots and scatter plots (Figures 7 and 8).

**Figure 7:**
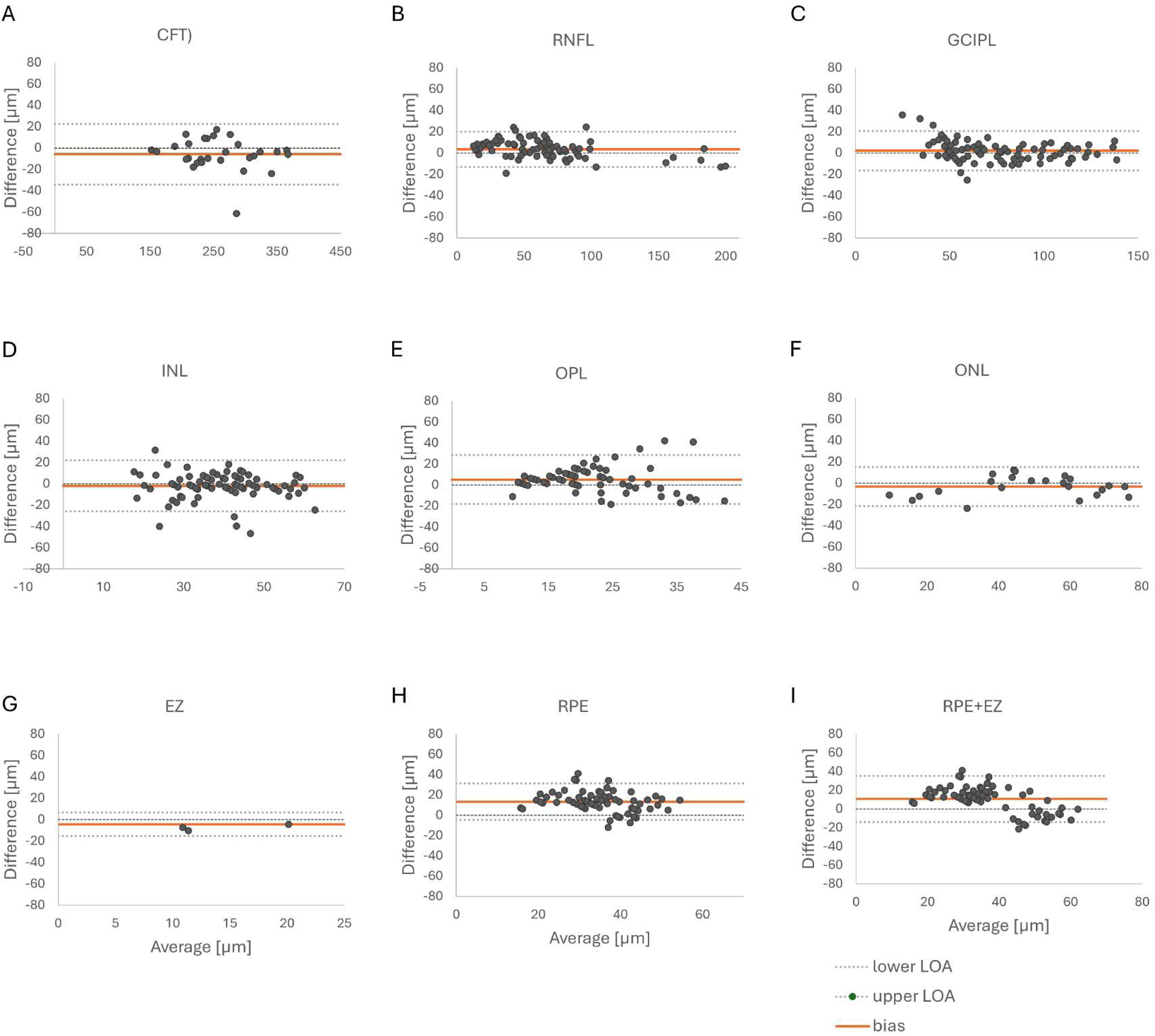
Bland-Altman Plots displaying agreement between OCT-SAM and manual measurements. The differences between measurements are plotted against their mean, with the mean difference (bias) and the upper and lower levels of agreement (LOA) shown.

**Figure 8:**
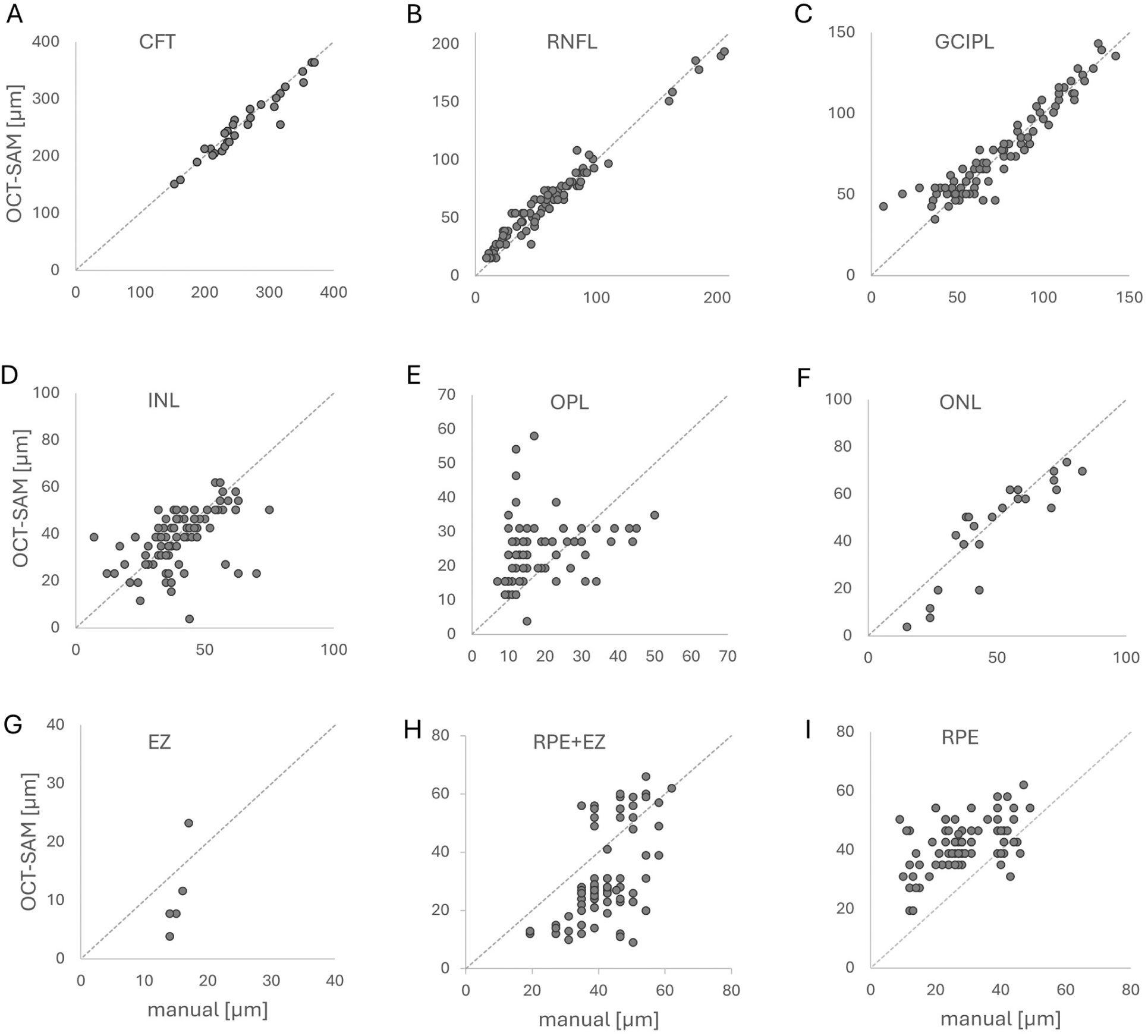
Scatter plots displaying OCT-SAM measurements against manual measurements (µm), with the line of identity indicated.

The expert grader detected the CFT, RNFL, GCIPL, INL and RPE in 90 out of 90 measurements, the OPL in 89, the ONL in 85, and the EZ in 21. OCT-SAM detected the CFT, RNFL, GCIPL, INL and RPE in all measurements, while detection dates were lower for OPL (72/90), ONL (23/90) and the EZ (6/90). We excluded the undetected layers for our subsequent calculation of mean and percentage differences. Overall, OCT-SAM demonstrated good agreement with manual measurements for CFT, RNFL and GCIPL, with relative differences ranging from 4.4% to 11.2%. Agreement of INL and ONL were lower, with relative differences of 21.8 and 22.5 %, respectively. Measurements of the EZ and RPE were least consistent, with relative differences of 54,8% and 41.1%, respectively (Table 11).

Agreement between automated and manual measurements was evaluated using Bland–Altman analysis (Figure 7, Table 12). We found minimal bias in the RNFL (3.4), ONL (−3.32), INL (−2.01) and GCIPL (2.01). Bias was slightly higher in the EZ (−4.36) and the CFT (−5.85), and largest in the RPE (13.39). As the detection rate of the EZ was low, we compared the bias of the sum of RPE and EZ with the OCT-SAM measurements, resulting in a slightly lower bias (10.38) compared to the RPE alone.

Overall, OCT-SAM showed accurate layer detection in the inner retina layers (RNFL, GCIPL, INL) with good agreement when compared to manual measurements. The outer retina layer (RNFL) was also detected accurately whilst the low detection rate of the EZ-layer resulted in inconsistent thickness measurements.

### OCT-SAM enables the efficient retrospective analysis of RP patients

We used a retrospective longitudinal dataset to demonstrate the clinical relevance of OCT-SAM for monitoring retinal layer degeneration as well as detecting pathological shifts in retinal layers of RP patients (Table 13). Three patients were selected who presented at our clinic for imaging diagnostic appointments at three different time points spanning a minimum of 3 years. RP01 was staged as early RP and showed clinically stable disease during the first two appointments with development of a CME at their last visit. RP57, staged as early RP, displayed a worsening CME over the course of the observed timespan. RP42 displayed an advanced stage of RP with clinically stable disease progression (Figure 9).

**Figure 9:**
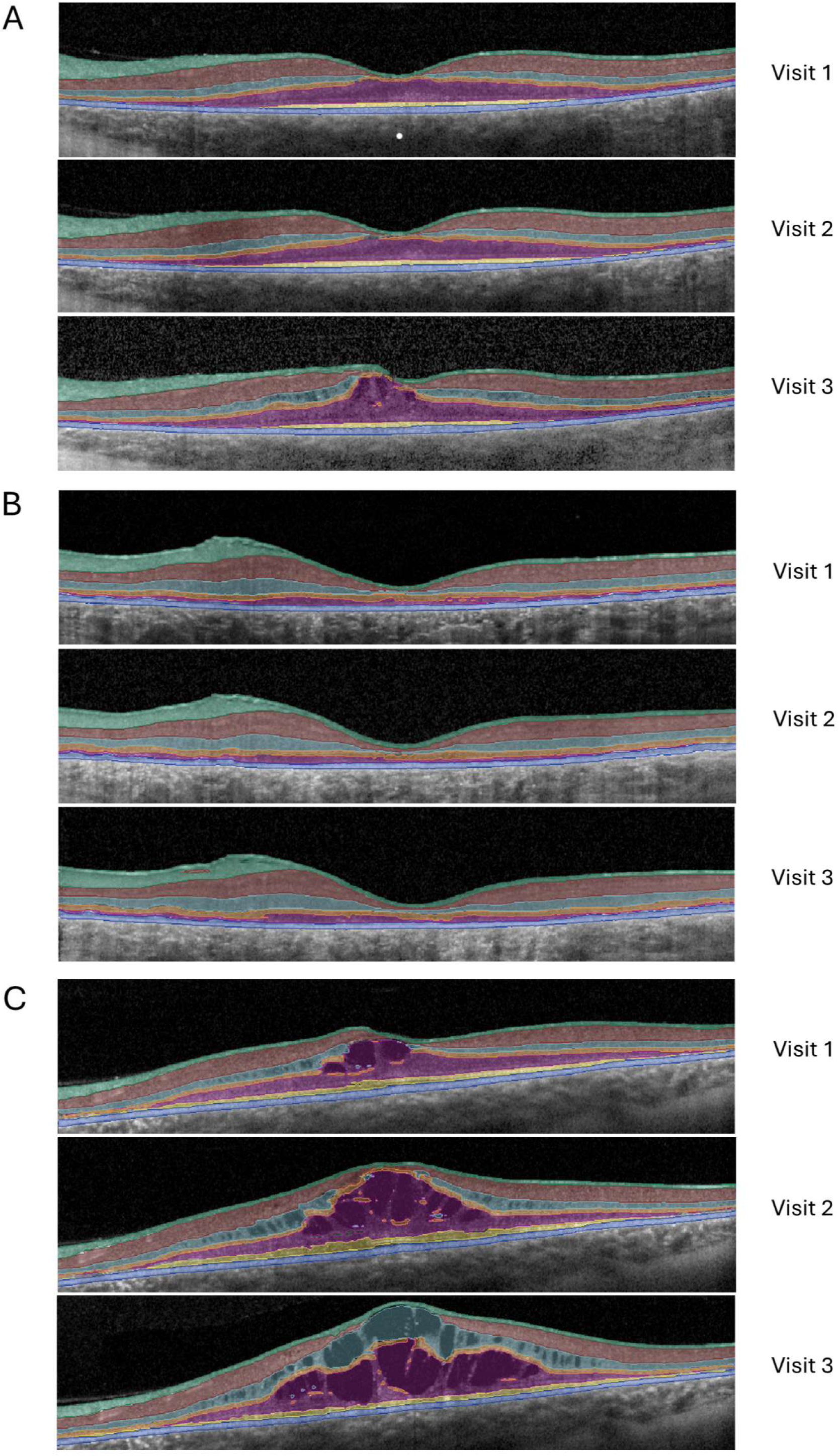
Interactively segmented OCTs over the course of three visits spanning 3 years. Row A: Patient RP01 shows stable progression with development of a CME in the last check-up. Row B: Patient RP42 shows overall stable disease progression. Row C: Patient RP57 presenting with an increasing cystoid macular edema over time. RNFL = turquoise, GCIPL = brown, INL = light blue, OPL = orange, ONL = purple, EZ = yellow, RPE = dark blue.

We selected the central B-Scan of the left eye for each visit and applied OCT-SAM’s integrated automated measurement tool for mean individual retinal layer thicknesses and central fovea thickness (CFT). Automated segmentations were refined using the napari-integrated interactive segmentation tool. A maximum of two positive point prompts per layer were applied to include layers missed by the automated segmentation while negative points were used to correct layer attribution in the presence of cystoid macular edema. No additional manual re-drawing or pixel-corrections were performed (Figure 10).

**Figure 10:**
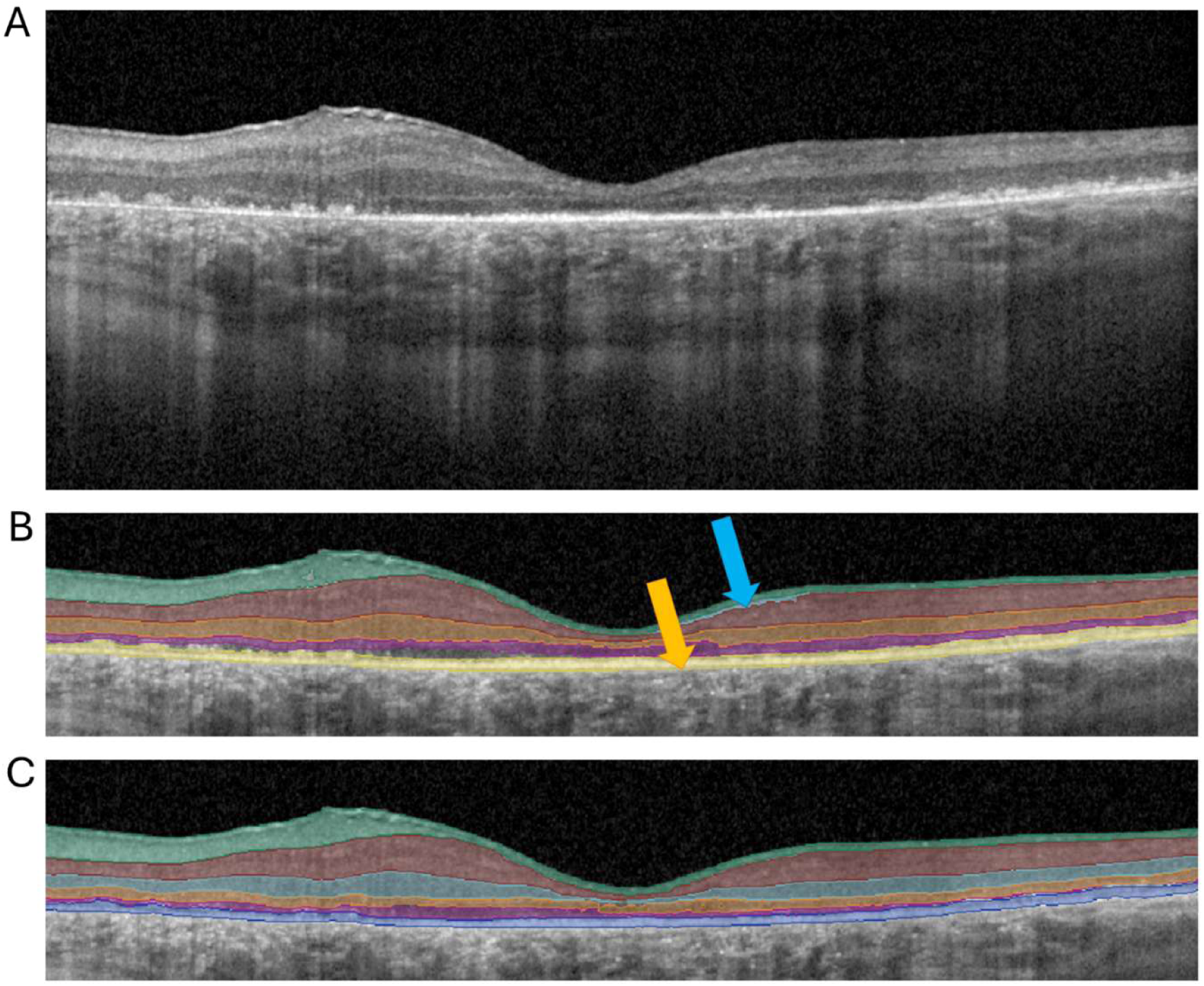
Process of interactive segmentation using OCT-SAM. Image A: raw OCT midline B-scan. Image B: automated model segmentation, masked. Blue arrow: additional layer found between RNFL and GCIPL. Yellow arrow: last layer segmented as EZ. C: segmentation after minor corrections via interactive annotation tool, masked.

To test whether general degeneration trends or pathologies could be identified by automated segmentation without interactive correction, we compared the automated and the corrected retinal layer thicknesses. We chose the CFT, RNFL, GCIPL, INL and RPE for this analysis, since OCT-SAM had demonstrated a 100% detection rate and accurate segmentation of these layers in our validation analysis. The RNFL, GCIPL and INL were defined as the first three consecutive layers identified by OCT-SAM, while the RPE was set as the last layer in each B-scan sequence.

Both automated and corrected measurements captured the same longitudinal trends across all three patients (Table 14, Figure 11). Increasing CFT values reflected progressive CME development in RP01 and RP57, whereas RP42 showed an overall reduction in CFT correlating with continued degenerative processes as part of the underlying disease. While the automated measurement slightly overestimated some measurements, both automated and corrected segmentations accurately reflected the same underlying trends and pathologies.

**Figure 11:**
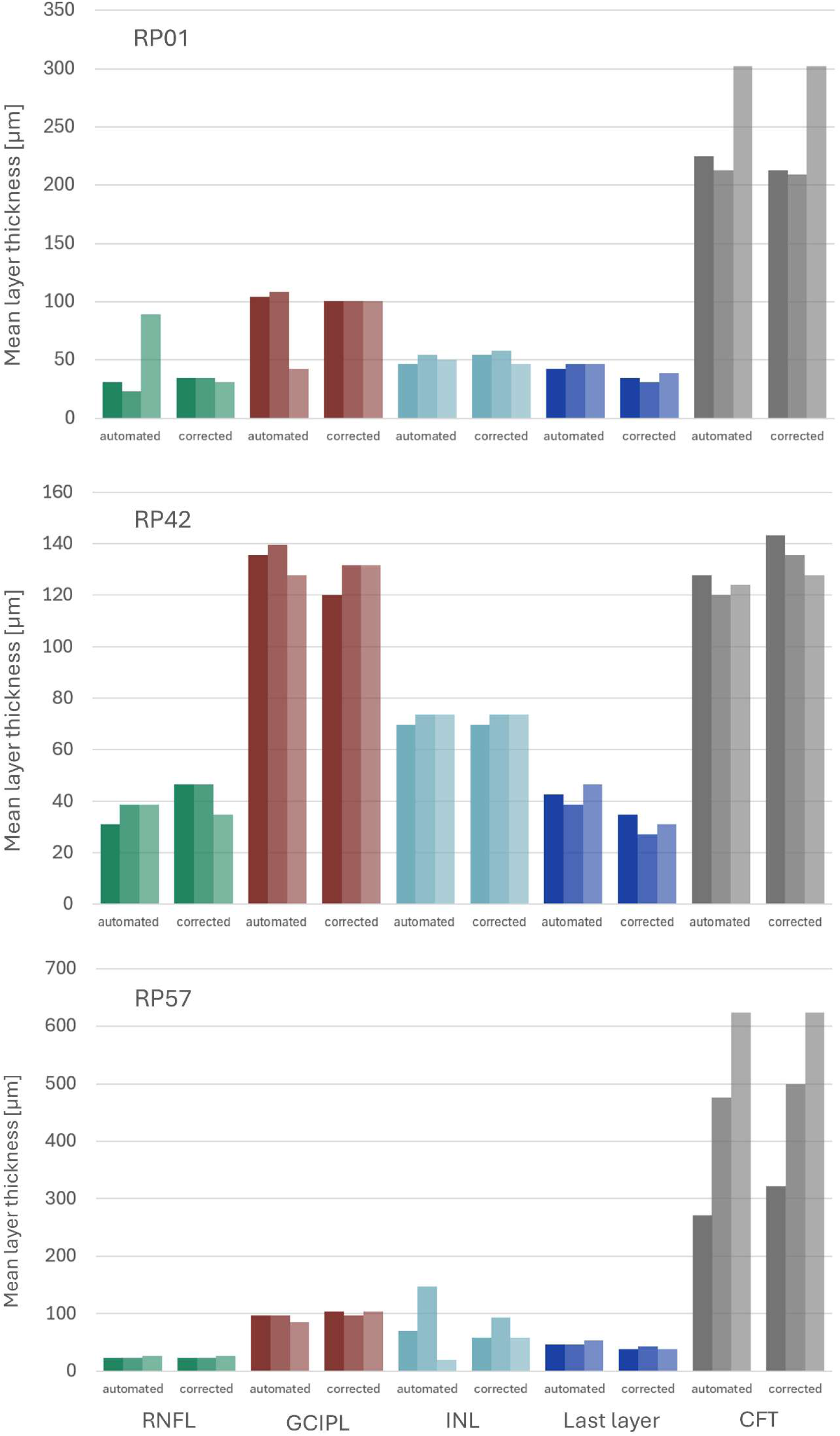
Bar plots of RNFL, GCIPL, INL, RPE (last layer) and CFT thicknesses in three patients over time with. Left –corrected segmentation, right – automated segmentation. Dark color = first visit, medium color = middle visit, light color = last visit.

This clinical implementation shows that OCT-SAM is able to detect existing and newly emerging pathologies such as CME in OCT scans. It also shows robust longitudinal reproducibility of measurements across time in absence of marked morphological changes, while detecting trends intrinsic to slow degenerative processes.

## Discussion

### Performance of OCT-SAM vs nnU-Net

In this study, we have demonstrated the accurate performance of two deep learning models to segment inner retinal layers in RP OCT-data. This was achieved with a comparatively small disease-specific dataset in combination with pretraining and post-processing strategies. Furthermore, we have introduced OCT-SAM’s interactive measurement tool enabling study specific quantification of longitudinal degenerative changes in RP retina.

Previous deep learning approaches in IRD and RP primarily focus on the segmentation of outer retina layers in OCT scans such as the EZ, OS (outer segment)-metrics, RPE, and ONL ^15,16^, all of which have been shown to correlate with functional parameters such as visual acuity ^26,27^ and visual field ^28,29^. A hybrid deep machine learning model of two convolutional neural networks, U-Net and a sliding window algorithm, was trained to segment OCT-images into four layers: ILM-dINL (distal inner nuclear layer), dINL-EZ, EZ-pRPE (proximal RPE), and pRPE-BM ^17^. For disease progression, a deep convolutional neural network model was trained to track the EZ-width as well as OS metrics RP-patients with a RPGR mutation ^30^. AIRDetect-OCT detects basal lamina (BL) to inner limiting membrane (ILM) retina boundaries, intraretinal cystic spaces, subretinal fluid, subretinal hyperreflective material, pigment epithelium detachment, EZ loss, RPE-loss and the fovea center ^31^. Lastly, a U-Net based model was trained to detect the ONL-layer ^13^.

To our knowledge, deep learning has not yet been applied to inner retina segmentation in RP. Existing cross-sectional and retrospective studies on inner retina health in RP suggest that inner retina layers remain relatively stable in early disease stages but show degenerative changes in succession with outer retinal layers in moderate and advanced RP ^5,32,33^.

To track disease progression over time, we have introduced two complementary models serving different purposes. Our first model, OCT-SAM, demonstrated accurate automated layer segmentation particularly of the RNFL, GCIPL, INL and the RPE. To our knowledge, OCT-SAM has not yet been used for retina layer segmentation. The integrated napari plugin enables efficient interactive correction and refinement of the automated segmentation via point and box prompts, including layers such as the OPL, ONL and the EZ. The napari interface further allows the integration of automated measurements tailored to specific study outcomes. We have integrated a “measure” tool, deriving the minimum, maximum and mean layer thicknesses as well as the CFT from segmented B-scans. Here, OCT-SAM achieved good agreement with manual measurements.

Additionally, OCT-SAM’s interactive segmentation tool was central to our training data generation, showcasing the effectiveness of combining interactive and automatic segmentation to accelerate the annotation process. For the second, third and fourth training cycle, OCT-data were annotated using the latest version of OCT-SAM and its integrated interactive annotation tool. Both automated and interactive segmentation improved across training cycles, reducing the need for manual corrections.

Our second model, nnU-Net, showed overall excellent segmentation performance for RP retinal layers. While OCT-SAM achieved a precision of 0.93, recall of 0.8, F-1 score of 0.85 and a dice score of 0.77, nnU-Net surpassed OCT-SAM in automated segmentation with a precision, recall and F-1 score of 0.96 and a dice-score of 0.88. In contrast to OCT-SAM, whose primary asset is the interactive annotation and study specific tool integration, nnU-Net enables precise, fully automated segmentation of RP datasets including accurate anatomical allocation via integrated semantic layer prediction.

Prior deep-learning based approaches for OCT-segmentation in IRDs rely on large training datasets for which openly available datasets are scarce ^13^ and image quality is often reduced due to fixation issues in advanced RP stages ^14^. RP or IRD datasets of prior studies have ranged from 480 manually annotated OCT-midline B-Scans of 220 RP patients ^17^, 1,749 annotated B-scans across 275 patients with a variety of IRDs ^31^, to 25 volume scans of 16 RP patients, each consisting of either 49 or 97 B-Scans ^13^. Here, we have used a combination of existing open source non-IRD datasets with a total of 1,700 annotated B-Scans to pre-train the model. The first provided training on non-specific retinal layer anatomy ^25^, while the second ^24^ enhanced the model’s robustness for CME detection, a common complication affecting up to 50% of RP patients ^34^. A similar approach was followed by Almushattat et al. ^35^, enabling accurate CME-quantification in RP patients. After pre-training, we included disease-specific training data consisting of a variety of RP stages and characteristic complications. The performance of both models plateaued at around 100 RP-images, demonstrating that a large disease-specific dataset is not necessary for excellent model performance. For postprocessing, we developed an algorithm that merges segmented regions and assigns them to anatomically plausible retinal layers based on anatomical priors. In contrast to previous post-processing approaches, which primarily focus on artifact removal and layer surface smoothing ^36^ or address layer boundary disruptions caused by degenerative pathologies via probability maps ^37,38^ or bilateral threshold distance maps ^39^, our algorithm uses semantic layer predictions which enforces anatomical consistency in the presence of retinal degenerative processes characteristic for RP. Through this three-step process, we have shown that adequate pre-training and disease-specific semantic post­processing can provide similar model performance results to previous IRD studies, reducing the necessity of disease-specific annotation and imaging data in a patient group where high-quality scans are rare.

In summary, while current automated segmentation algorithms fail to correctly segment RP because of disease characteristic degenerative processes ^6,9^, our models enable accurate monitoring and automated measurements of RP inner retinal layers without the need for time-consuming manual segmentations. We have performed all further evaluations with our OCT-SAM model because it provides both interactive and automatic segmentation functionality and is integrated with a user-friendly tool. We have made this tool as well as both models and their training routines publicly available for use by the community.

### Clinical application of OCT-SAM

As new forms of treatment such as optogenetics show promising advances in clinical trials ^3^, monitoring and quantifying inner retina health in RP has increased in clinical importance. In contrast to gene therapy, optogenetics is a mutation-independent approach targeting neurons of the inner retina: RGCs and BPCs are stimulated to express light-sensitive opsins, superseding the function of photoreceptor cells ^40^. Results from recent clinical trials show a sustained vision gain under treatment with a multi-characteristic opsin targeting BPCs ^41^. Emerging low light sensitive opsins, e.g. ChReef, demonstrate effective vision restoration when targeting RGCs in a RP mouse model^42^. To translate this promising therapeutic approach into clinical use for RP patients, precise measurement of retinal layer degeneration is necessary to ensure optimal patient selection, as optogenetics targeting RGCs requires a largely intact inner retina. At the same time, the reliable assessment and segmentation of retinal layers in OCT scans of RP patients is often compromised by the gradual loss of the outer layers and the frequent occurrence of CME. To demonstrate that OCT-SAM has the potential to overcome these obstacles, we implemented OCT-SAM as a longitudinal monitoring tool to track retinal layer changes in three RP patients. The time of observation spanned three years. Using the integrated measurement tool, we derived automated and corrected mean thicknesses of the RNFL, GCIPL, INL, RPE and the CFT. OCT-SAM enabled monitoring of overall stable inner retinal layers in early-staged RP (RP01) while detecting newly emerging pathological changes such as CME via increased CFT-values. Similarly, inner retinal layers of a second early-stage RP patient (RP57) remained relatively constant while a worsening CME over time was reflected by a steadily increasing CFT. In contrast, an advanced case of RP (RP42) showed reduced mean thicknesses of both outer and inner retinal layers, i.e. the RPE, the CFT and the RNFL. These results are consistent with current findings in literature, in which inner retinal layers remain largely intact in early RP stages while a thinning of the RNFL has been observed in advanced RP ^5,33^. Additionally, mean inner retina thicknesses remained relatively stable overall and may be less affected by CME development, a complication found commonly in RP^34^. Overall, OCT-SAM provided effective tracking of degenerative changes and enabled the detection of newly emerging pathologies such as CME both in automated and corrected output.

### Limitations and future perspectives

Working with a smaller dataset entails that not every mutation variant of RP and correlating intrinsic degenerative differences between genotypes might be accounted for in the training data. While inner retina layers were accurately detected, OCT-SAM demonstrated inconsistent performance for outer retina layers, specifically the ONL and EZ-layer, central to RP disease staging.^23^

We intend to address this limitation in our future work, where we plan to expand 2D-annotation of OCT-SAM 1.0 to 3D-annotation, thus integrating more consistent semantic layer assignment and degenerative pattern predictions across OCT volume scans.

## Data Availability

All data produced in the present work are contained in the manuscript and available online at https://github.com/computational-cell-analytics/oct-sam.

## Acknowledgments

This work was supported by the German Research Foundation (Deutsche Forschungsgemeinschaft, DFG) through grants SFB1690/B06 (to E.M.) and SFB1690/A09 (to C.P.). The DFG funds M.S. under Germany’s Excellence Strategy—EXC 2067/1-390729940. D.L., E.M., S.B., and C.P. were supported by the Else Kröner Fresenius Foundation and zukunft.niedersachsen, the joint science funding program of the Lower Saxony Ministry of Science and Culture and the Volkswagen Foundation, via the Else Kröner Fresenius Center for Optogenetic Therapies” (EKFZ). C.P. is also supported by the Ministry of Science and Culture of Lower Saxony through funds from the program zukunft.niedersachsen of the Volkswagen Foundation for the “CAIMed –Lower Saxony Center for Artificial Intelligence and Causal Methods in Medicine” project (grant no. ZN4257).

We also gratefully acknowledge the computing time granted by the Resource Allocation Board and provided on the supercomputer Lise and Emmy at NHR@ZIB and NHR@Göttingen as part of the NHR infrastructure. The calculations for this research were conducted with computing resources under the project nim00007.

## Tables

**1.** Dataset for first training cycle and validation test:

| PS | Age range | Associated RP gene | RP-stage OD | complications OD | RP-stage OS | complications OS |
| --- | --- | --- | --- | --- | --- | --- |
| RP01 | 30-34 | USH2A | early | CME, gliosis | early | CME, gliosis |
| RP04 | 30-34 | RPGR | moderate | gliosis, RIQ | advanced | gliosis, RIQ |
| RP07 | 35-39 | CNGB1, RDH11 | early | none | early | none |
| RP13 | 20-24 | RHO | early | CME, gliosis | early | CME, gliosis |
| RP14 | 30-34 | EYS, PRPF3 | moderate | gliosis | moderate | gliosis |
| RP15 | 50-54 | p.a. | moderate | CME | moderate | CME |
| RP16 | 50-54 | PDE6B | advanced | CME | advanced | CME |
| RP17 | 25-29 | FLVCR1 | moderate | CME, CME | moderate | CME |
| RP19 | 65-69 | USH2A | early | none | early | gliosis |
| RP20 | 20-24 | PDE6A | early | CME, gliosis | moderate | CME, gliosis |
| RP21 | 75-79 | p.a. | early | CME, gliosis | early | none |
| RP23 | 25-29 | PDE6A | moderate | CME, RIQ | moderate | CME, RIQ |

PS = pseudonym, YOB = year of birth, RIQ = reduced image quality of OCT scan, OD = right eye, OS = left eye, p.a. = pending analysis

**2.** Dataset for second training cycle:

2. Dataset for second training cycle:
| PS | Age range | Associated RP gene | RP-stage OD | complications OD | RP-stage OS | complications OS |
| --- | --- | --- | --- | --- | --- | --- |
| RP19 | 65-69 | USH2A | early | none | early | gliosis |
| RP41 | 30-34 | USH2A | moderate | none | moderate | gliosis |
| RP58 | 35-39 | USH2A | early | RIQ | early | CME, gliosis |
| RP101 | 15-19 | p.a. | early | RIQ, gliosis | - | - |
| RP122 | 35-39 | RPGR | advanced | RIQ, gliosis | advanced | gliosis |

**3.** Dataset for third training cycle:

3. Dataset for third training cycle:
| PS | Age range | Associated RP gene | RP-stage OD | complications OD | RP-stage OS | complications OS |
| --- | --- | --- | --- | --- | --- | --- |
| RP27 | 65-69 | EYS, RP2 | early | RIQ | - | - |
| RP37 | 55-59 | CRX | moderate | gliosis | - | - |
| RP45 | 15-19 | MYO7A | moderate | CME | moderate | CME |
| RP46 | 30-34 | p.a. | moderate | gliosis | moderate | gliosis |
| RP49 | 50-54 | PCARE | moderate | central atrophy | - | - |
| RP101 | 15-19 | p.a. | - | - | early | RIQ, gliosis |
| RP254 | 5-9 | USH2A | moderate | gliosis | - | - |

**4.**
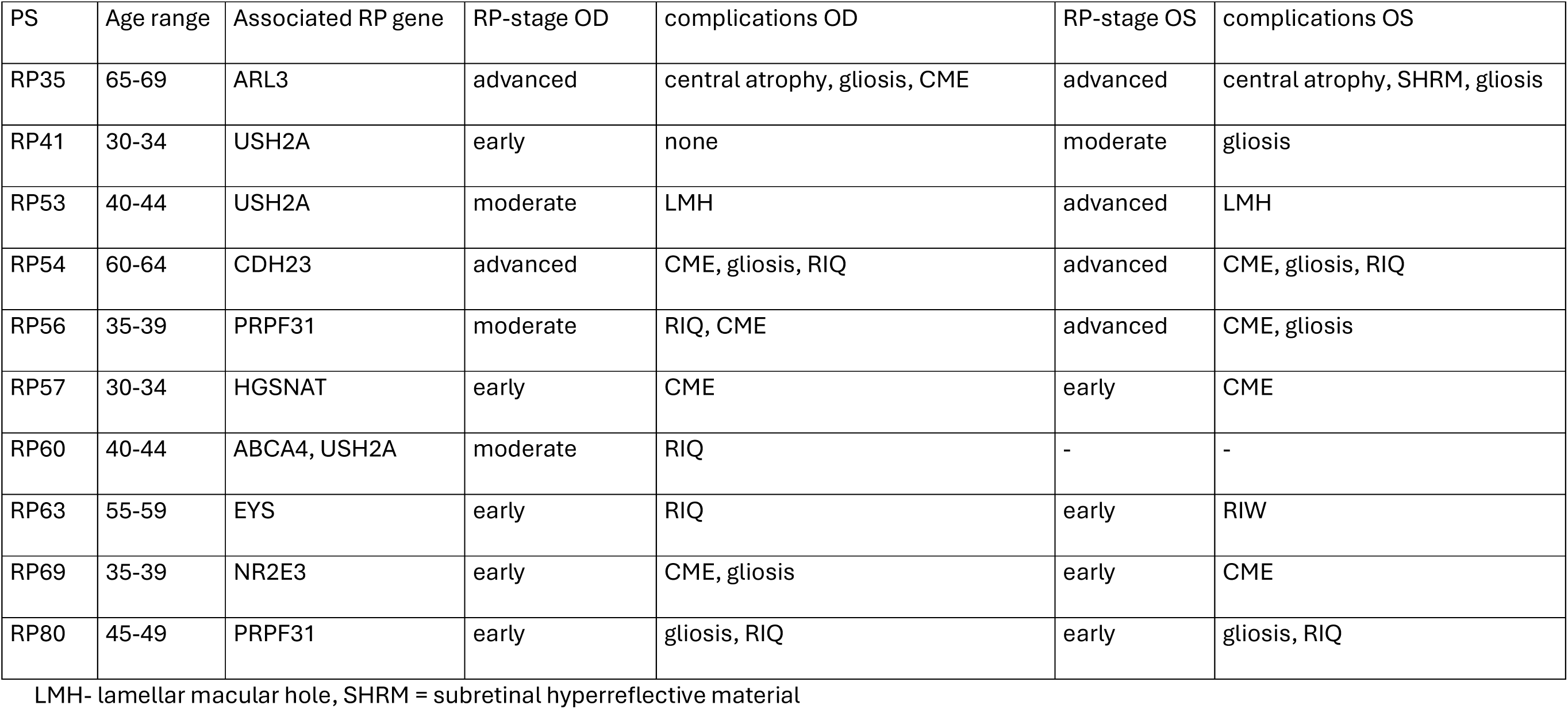
Dataset for fourth training cycle:

## Tables 5-8: Selected OCT-B-scans from 49-B-scan SD-OCT for annotation (z0 = first B-scan, z48 = last B-scan of 615 49-B-scan SD-OCT)

**5.**
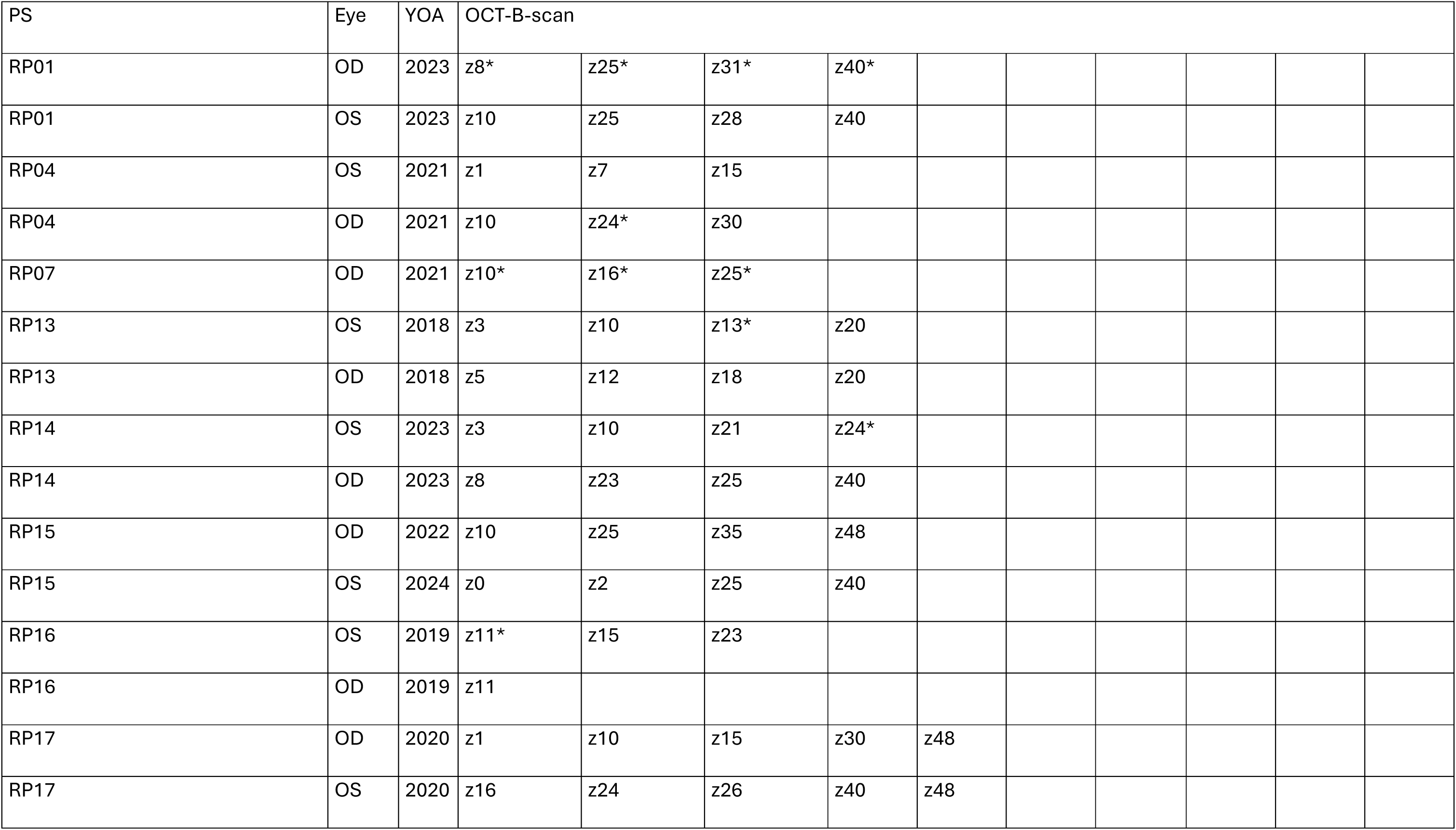
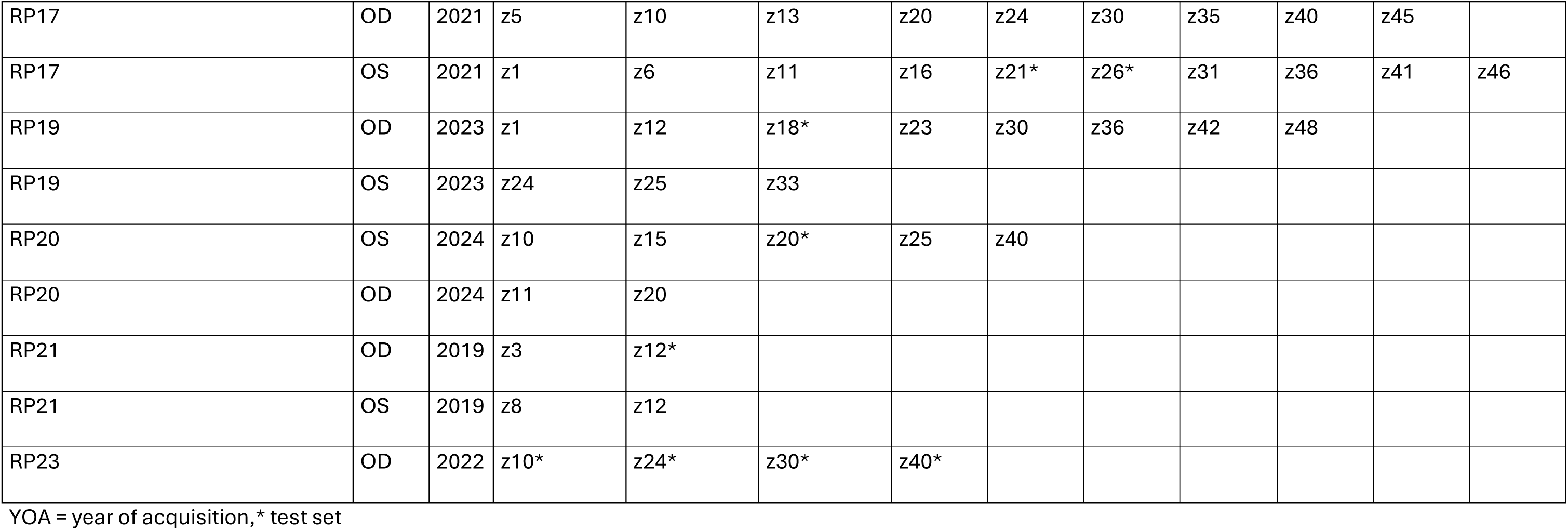
First training cycle and test set.

LMH- lamellar macular hole, SHRM = subretinal hyperreflective material 5. First training cycle and test set
| PS | Eye | YOA | OCT-B-scan |  |  |  |  |
| --- | --- | --- | --- | --- | --- | --- | --- |
| RP01 | OD | 2023 | z8* | z25* | z31* | z40* |  |
| RP01 | OS | 2023 | z10 | z25 | z28 | z40 |  |
| RP04 | OS | 2021 | z1 | z7 | z15 |  |  |
| RP04 | OD | 2021 | z10 | z24* | z30 |  |  |
| RP07 | OD | 2021 | z10* | z16* | z25* |  |  |
| RP13 | OS | 2018 | z3 | z10 | z13* | z20 |  |
| RP13 | OD | 2018 | z5 | z12 | z18 | z20 |  |
| RP14 | OS | 2023 | z3 | z10 | z21 | z24* |  |
| RP14 | OD | 2023 | z8 | z23 | z25 | z40 |  |
| RP15 | OD | 2022 | z10 | z25 | z35 | z48 |  |
| RP15 | OS | 2024 | z0 | z2 | z25 | z40 |  |
| RP16 | OS | 2019 | z11* | z15 | z23 |  |  |
| RP16 | OD | 2019 | z11 |  |  |  |  |
| RP17 | OD | 2020 | z1 | z10 | z15 | z30 | z48 |
| RP17 | OS | 2020 | z16 | z24 | z26 | z40 | z48 |

|  |  |  |  |  |  |  |  |  |  |  |  |  |
| --- | --- | --- | --- | --- | --- | --- | --- | --- | --- | --- | --- | --- |
| RP17 | OD | 2021 | z5 | z10 | z13 | z20 | z24 | z30 | z35 | z40 | z45 |  |
| RP17 | OS | 2021 | z1 | z6 | z11 | z16 | z21* | z26* | z31 | z36 | z41 | z46 |
| RP19 | OD | 2023 | z1 | z12 | z18* | z23 | z30 | z36 | z42 | z48 |  |  |
| RP19 | OS | 2023 | z24 | z25 | z33 |  |  |  |  |  |  |  |
| RP20 | OS | 2024 | z10 | z15 | z20* | z25 | z40 |  |  |  |  |  |
| RP20 | OD | 2024 | z11 | z20 |  |  |  |  |  |  |  |  |
| RP21 | OD | 2019 | z3 | z12* |  |  |  |  |  |  |  |  |
| RP21 | OS | 2019 | z8 | z12 |  |  |  |  |  |  |  |  |
| RP23 | OD | 2022 | z10* | z24* | z30* | z40* |  |  |  |  |  |  |

YOA = year of acquisition, \* test set

**6.** Second training cycle.

6. Second training cycle
| PS | Eye | YOA | Selected OCT-B-scans for annotation |  |  |
| --- | --- | --- | --- | --- | --- |
| RP19 | OD | 2025 | z10 | z25 | z27 |
| RP19 | OS | 2025 | z5 | z25 | z30 |
| RP122 | OD | 2025 | z8 | z26 | z48 |
| RP122 | OS | 2025 | z8 | z25 | z48 |
| RP41 | OD | 2025 | z10 | z25 | z27 |
| RP41 | OS | 2025 | z22 | z25 | z40 |
| RP58 | OD | 2025 | z5 | z25 |  |
| RP58 | OS | 2025 | z5 | z25 | z30 |
| RP101 | OD | 2025 | z5 | z25 | z30 |

**7.** Third training cycle.

7. Third training cycle
| PS | Eye | YOA | Selected OCT-B-scans for annotation |  |  |
| --- | --- | --- | --- | --- | --- |
| RP27 | OD | 2021 | z10 |  |  |
| RP37 | OD | 2021 | z10 | z25 | z30 |
| RP45 | OD | 2025 | z5 | z25 | z32 |
| RP45 | OS | 2025 | z5 | z25 | z32 |
| RP46 | OD | 2021 | z2 | z25 | z30 |
| RP46 | OS | 2021 | z22 | z25 | z42 |
| RP49 | OD | 2022 | z2 | z25 | z32 |
| RP101 | OS | 2025 | z10 | z15 | z20 |
| RP254 | OD | 2025 | z4 | z25 | z30 |

**8.** Fourth training cycle.

8. Fourth training cycle
| PS | Eye | YOA | Selected OCT-B-scans for annotation |  |  |
| --- | --- | --- | --- | --- | --- |
| RP35 | OD | 2021 | z4 | z25 | z40 |
| RP35 | OS | 2021 | z4 | z25 | z40 |
| RP35 | OS | 2025 | z24 | z30 |  |
| RP35 | OD | 2025 | z24 | z30 |  |
| RP41 | OS | 2022 | z4 | z25 | z40 |
| RP41 | OD | 2022 | z4 | z25 | z40 |
| RP41 | OD | 2025 | z10 | z24 | z30 |
| RP41 | OS | 2025 | z10 | z24 | z30 |
| RP53 | OS | 2022 | z5 | z45 | z48 |
| RP53 | OD | 2022 | z5 | z40 | z47 |
| RP53 | OD | 2025 | z5 | z10 | z35 |
| RP54 | OD | 2022 | z10 | z15 | z20 |
| RP54 | OS | 2022 | z10 | z15 | z20 |
| RP54 | OD | 2024 | z5 | z24 | z42 |
| RP54 | OS | 2024 | z5 | z24 | z42 |
| RP56 | OD | 2022 | z15 | z40 |  |
| RP56 | OS | 2022 | z15 | z25 | z40 |
| RP56 | OD | 2025 | z4 | z24 | z30 |
| RP57 | OD | 2022 | z15 | z25 | z40 |
| RP57 | OS | 2022 | z15 | z25 | z40 |
| RP57 | OS | 2025 | z5 | z24 | z30 |
| RP57 | OD | 2025 | z5 | z24 | z30 |
| RP60 | OD | 2021 | z10 | z24 | z30 |
| RP63 | OD | 2023 | z10 | z24 | z30 |
| RP63 | OS | 2023 | z10 | z24 | z30 |
| RP63 | OD | 2024 | z20 | z25 | z45 |
| RP63 | OS | 2024 | z20 | z25 | z45 |
| RP69 | OS | 2023 | z5 | z25 | z48 |
| RP69 | OD | 2023 | z5 | z25 | z48 |
| RP69 | OD | 2024 | z10 | z24 | z30 |
| RP69 | OS | 2024 | z10 | z24 | z30 |
| RP80 | OS | 2024 | z10 | z25 | z30 |
| RP80 | OD | 2024 | z5 | z25 | z40 |
| RP80 | OS | 2025 | z10 | z24 | z30 |
| RP80 | OD | 2025 | z5 | z25 | z48 |

**9.** Dataset for measurement validation.

9. Dataset for measurement validation
| PS | Age range | Associated RP gene | RP-stage OD | complications OD | RP-stage OS | complications OS |
| --- | --- | --- | --- | --- | --- | --- |
| RP06 | 45-49 | USH2A | - | - | early | gliosis |
| RP09 | 50-54 | SNRNP200 | - | - | moderate | RIQ |
| RP22 | 35-39 | p.a. | early | CME, gliosis | - | - |
| RP26 | 60-64 | RHO | advanced | central atrophy,<br>SHRM | - | - |
| RP40 | 30-34 | EYS | - | - | early | CME, gliosis |
| RP225 | 25-29 | EYS | advanced | gliosis | - | - |

**10.**
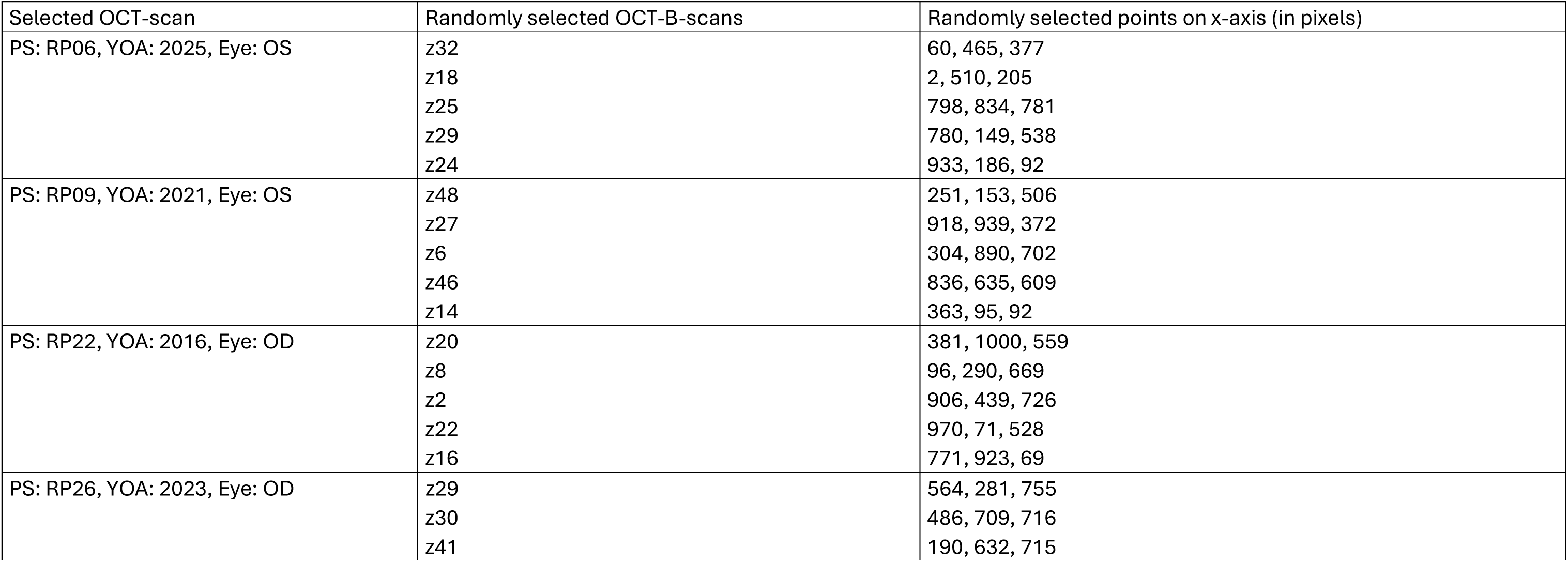
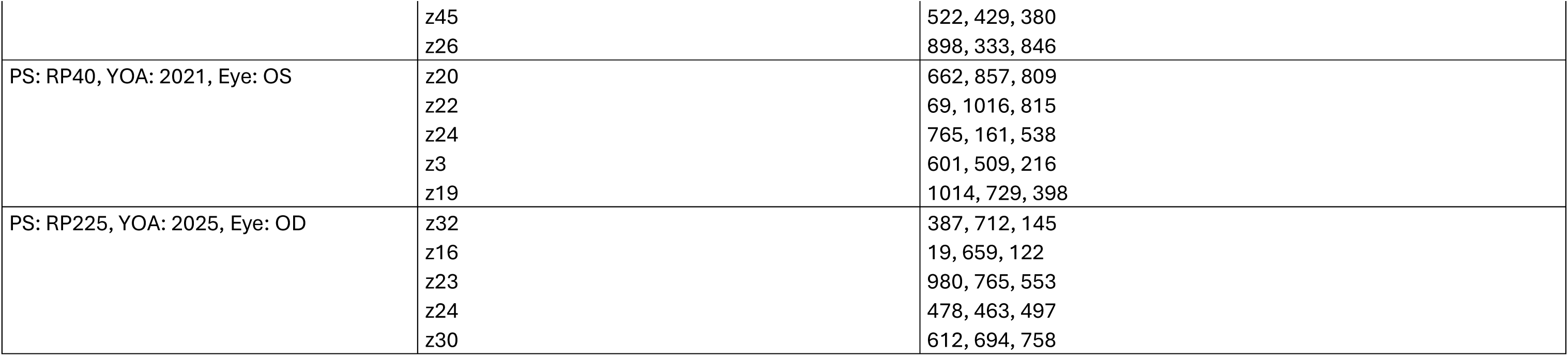
Selected B-scans, points for measurement validation.

**11.**
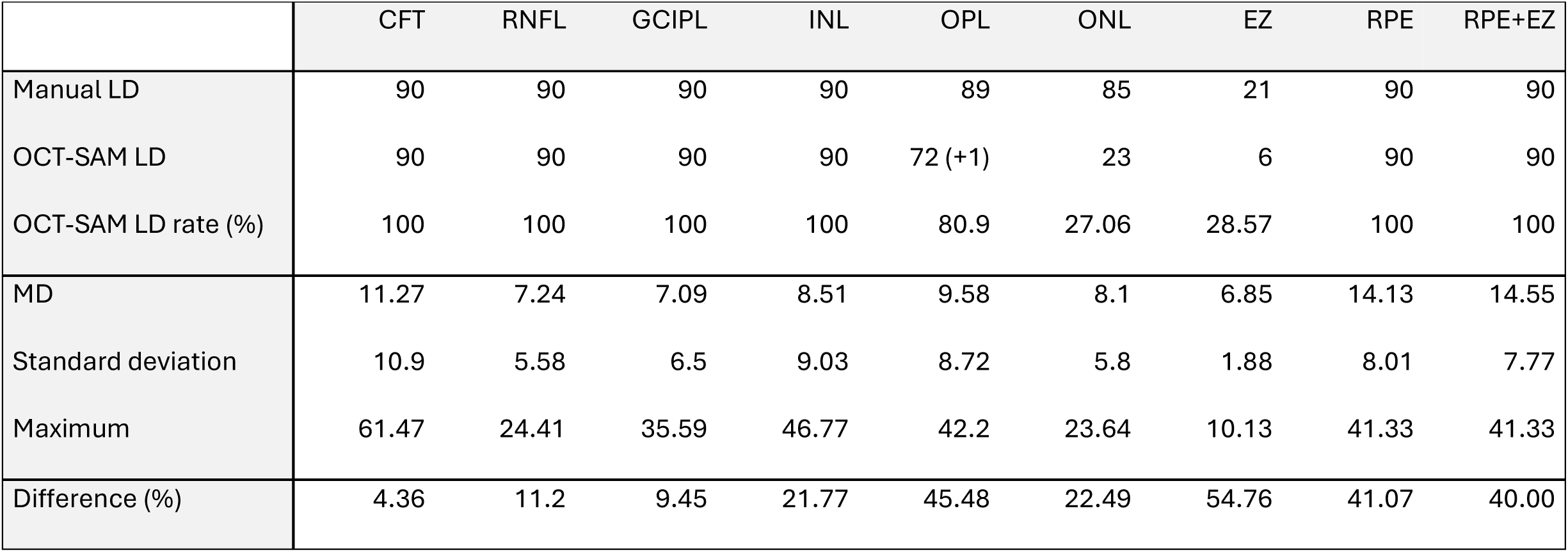
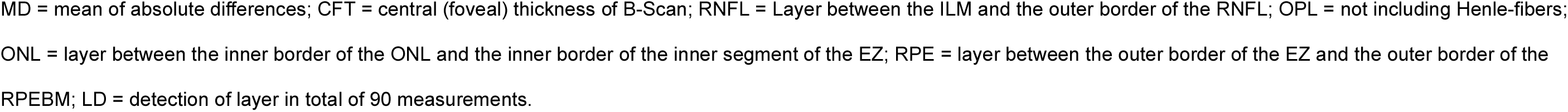
Comparison of manual and automated measurements: layer detection rate, absolute difference, standard deviation and percentage differences.

|  | CFT | RNFL | GCIPL | INL | OPL | ONL | EZ | RPE | RPE+EZ |
| --- | --- | --- | --- | --- | --- | --- | --- | --- | --- |
| Manual LD | 90 | 90 | 90 | 90 | 89 | 85 | 21 | 90 | 90 |
| OCT-SAM LD | 90 | 90 | 90 | 90 | 72 (+1) | 23 | 6 | 90 | 90 |
| OCT-SAM LD rate (%) | 100 | 100 | 100 | 100 | 80.9 | 27.06 | 28.57 | 100 | 100 |
| MD | 11.27 | 7.24 | 7.09 | 8.51 | 9.58 | 8.1 | 6.85 | 14.13 | 14.55 |
| Standard deviation | 10.9 | 5.58 | 6.5 | 9.03 | 8.72 | 5.8 | 1.88 | 8.01 | 7.77 |
| Maximum | 61.47 | 24.41 | 35.59 | 46.77 | 42.2 | 23.64 | 10.13 | 41.33 | 41.33 |
| Difference (%) | 4.36 | 11.2 | 9.45 | 21.77 | 45.48 | 22.49 | 54.76 | 41.07 | 40.00 |

MD = mean of absolute differences; CFT = central (foveal) thickness of B-Scan; RNFL = Layer between the ILM and the outer border of the RNFL; OPL = not including Henle-fibers; ONL = layer between the inner border of the ONL and the inner border of the inner segment of the EZ; RPE = layer between the outer border of the EZ and the outer border of the RPEBM; LD = detection of layer in total of 90 measurements.

**12.** Agreement between automated and manual measurements.

12. Agreement between automated and manual measurements
|  | CFT | RNFL | GCIPL | INL | OPL | ONL | EZ | RPE | RPE+EZ |
| --- | --- | --- | --- | --- | --- | --- | --- | --- | --- |
| Bias | -5.85 | 3.40 | 2.01 | -2.01 | 5.20 | -3.32 | -4.36 | 13.39 | 10.68 |
| Standard deviation | 14.55 | 8.49 | 9.40 | 12.25 | 11.86 | 9.39 | 5.61 | 9.20 | 12.57 |
| lower LOA | -34.36 | -13.23 | -16.42 | -26.01 | -18.05 | -21.74 | -15.36 | -4.64 | -13.95 |
| upper LOA | 22.66 | 20.03 | 20.45 | 21.99 | 28.45 | 15.09 | 6.64 | 31.43 | 35.31 |

Agreement is assessed via bias (mean differences in µm between the automated and manual measurements for identified retinal layers), standard deviation and levels of agreement. LOA = level of agreement

**13.** Dataset for clinical implementation of OCT-SAM.

| PS | Age range | Associated RP gene | RP-stage OS | complications OS |
| --- | --- | --- | --- | --- |
| RP01 | 30-34 | USH2A | early | CME, gliosis |
| RP42 | 25-29 | PRPH2 | late | none |
| RP57 | 30-34 | HGSNAT | early | CME |

**14.**
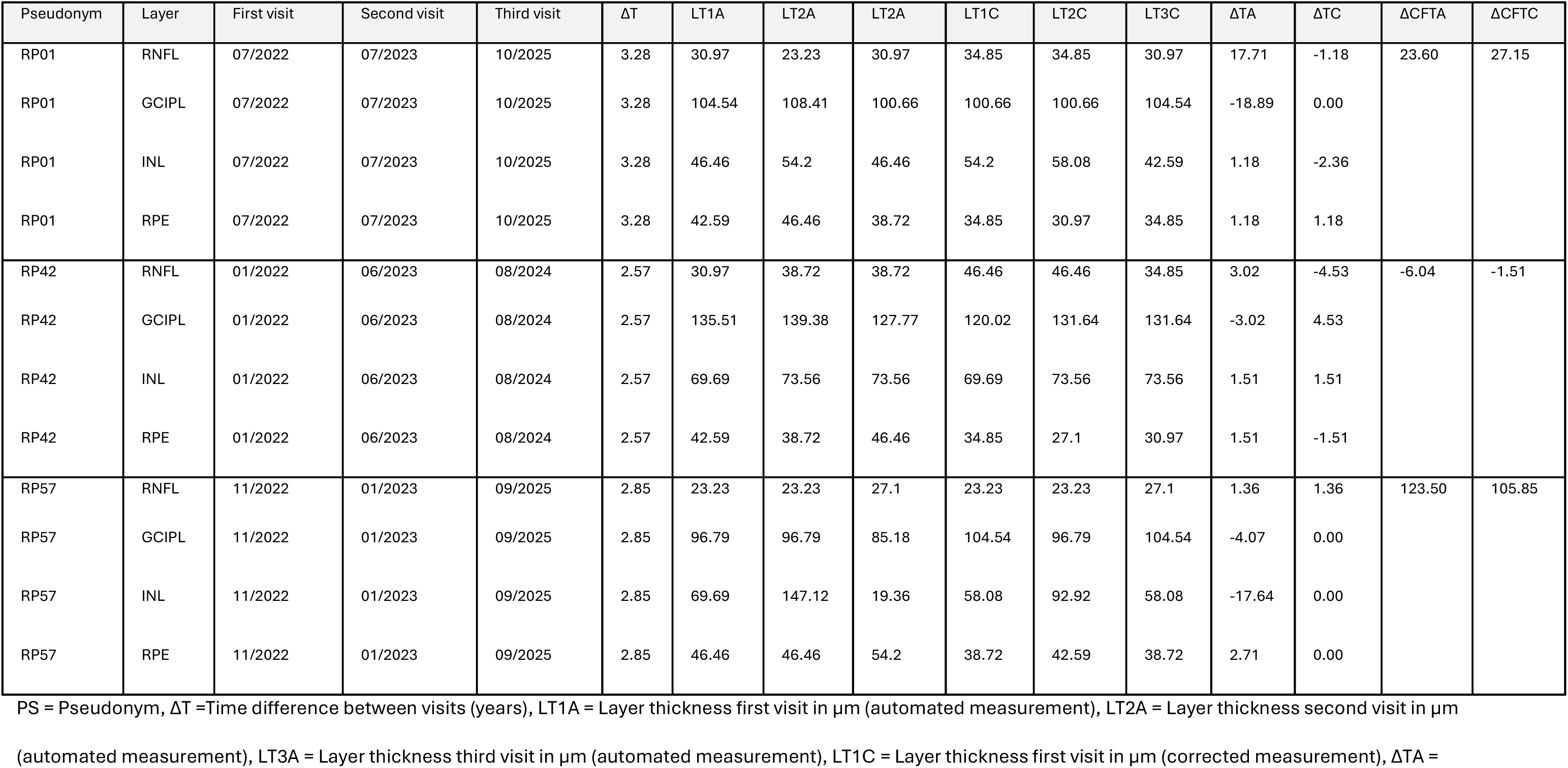
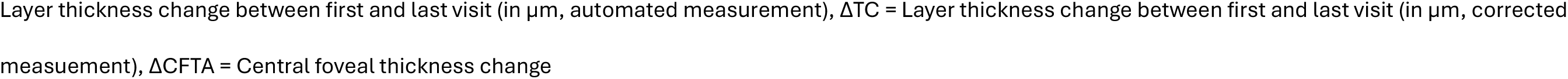
Longitudinal changes in mean layer thicknesses in three RP patients across three visits.

| Pseudonym | Layer | First visit | Second visit | Third visit | ΔT | LT1A | LT2A | LT2A | LT1C | LT2C | LT3C | ΔTA | ΔTC | ΔCFTA | ΔCFTC |
| --- | --- | --- | --- | --- | --- | --- | --- | --- | --- | --- | --- | --- | --- | --- | --- |
| RP01 | RNFL | 07/2022 | 07/2023 | 10/2025 | 3.28 | 30.97 | 23.23 | 30.97 | 34.85 | 34.85 | 30.97 | 17.71 | -1.18 | 23.60 | 27.15 |
| RP01 | GCIPL | 07/2022 | 07/2023 | 10/2025 | 3.28 | 104.54 | 108.41 | 100.66 | 100.66 | 100.66 | 104.54 | -18.89 | 0.00 |  |  |
| RP01 | INL | 07/2022 | 07/2023 | 10/2025 | 3.28 | 46.46 | 54.2 | 46.46 | 54.2 | 58.08 | 42.59 | 1.18 | -2.36 |  |  |
| RP01 | RPE | 07/2022 | 07/2023 | 10/2025 | 3.28 | 42.59 | 46.46 | 38.72 | 34.85 | 30.97 | 34.85 | 1.18 | 1.18 |  |  |
| RP42 | RNFL | 01/2022 | 06/2023 | 08/2024 | 2.57 | 30.97 | 38.72 | 38.72 | 46.46 | 46.46 | 34.85 | 3.02 | -4.53 | -6.04 | -1.51 |
| RP42 | GCIPL | 01/2022 | 06/2023 | 08/2024 | 2.57 | 135.51 | 139.38 | 127.77 | 120.02 | 131.64 | 131.64 | -3.02 | 4.53 |  |  |
| RP42 | INL | 01/2022 | 06/2023 | 08/2024 | 2.57 | 69.69 | 73.56 | 73.56 | 69.69 | 73.56 | 73.56 | 1.51 | 1.51 |  |  |
| RP42 | RPE | 01/2022 | 06/2023 | 08/2024 | 2.57 | 42.59 | 38.72 | 46.46 | 34.85 | 27.1 | 30.97 | 1.51 | -1.51 |  |  |
| RP57 | RNFL | 11/2022 | 01/2023 | 09/2025 | 2.85 | 23.23 | 23.23 | 27.1 | 23.23 | 23.23 | 27.1 | 1.36 | 1.36 | 123.50 | 105.85 |
| RP57 | GCIPL | 11/2022 | 01/2023 | 09/2025 | 2.85 | 96.79 | 96.79 | 85.18 | 104.54 | 96.79 | 104.54 | -4.07 | 0.00 |  |  |
| RP57 | INL | 11/2022 | 01/2023 | 09/2025 | 2.85 | 69.69 | 147.12 | 19.36 | 58.08 | 92.92 | 58.08 | -17.64 | 0.00 |  |  |
| RP57 | RPE | 11/2022 | 01/2023 | 09/2025 | 2.85 | 46.46 | 46.46 | 54.2 | 38.72 | 42.59 | 38.72 | 2.71 | 0.00 |  |  |

PS = Pseudonym, ΔT =Time difference between visits (years), LT1A = Layer thickness first visit in μm (automated measurement), LT2A = Layer thickness second visit in μm
(automated measurement), LT3A = Layer thickness third visit in μm (automated measurement), LT1C = Layer thickness first visit in μm (corrected measurement), ΔTA =
Layer thickness change between first and last visit (in $\mu\text{m}$ , automated measurement), $\Delta\text{TC}$ = Layer thickness change between first and last visit (in $\mu\text{m}$ , corrected measuement), $\Delta\text{CFTA}$ = Central foveal thickness change

